# Differential Associations of Heat Metrics with Health Statistics: A Spatiotemporal Analysis of Temperature Indicators and Mortality Across Thailand Provinces

**DOI:** 10.1101/2025.10.01.25337088

**Authors:** Jacques Wels, Romnalin Keanjoom, Juan Gonzalez Hijon, Sittichai Choosumrong

## Abstract

**Background:** Assessments of climate change health impacts often rely on ambient temperature, often neglecting the role of humidity and wind speed. This can be problematic in countries with high climate diversity and pronounced climate warming as it is the case in many South-East Asian nations. This study compared trends in multiple heat metrics (i.e., actual heat, Heat Index, Humidex and Apparent Temperature) across Thailand’s provinces and evaluated their respective associations with province-level mortality.

**Methods:** We analysed daily meteorological data (2006-2024) to calculate trends across heat metrics for 71 provinces (6 missing). Then, using negative binomial mixed-effects regression models, we assessed the 1-year lagged associations between each annual heat indicator and all-cause mortality from 2008 to 2023, adjusting for year and province.

**Results:** Composite heat indices increased at a faster rate than ambient temperature in the majority of provinces. All composite heat metrics surpassed actual heat in predicting mortality. Specifically, our analyses revealed that a 1°C increase in one-year lagged Apparent Temperature was associated with 47,196 Excess Deaths (ED) (95% CI: 13,287 to 82,704) over the study period, representing the strongest association among the metrics tested. However, the strength of the association varied across regions with different climate trajectories. Stratified analyses by death in- and out-of-hospital settings show strong association of Humidex and Apparent Temperature with hospital deaths at 1-year lag but out-of-hospital deaths show no associations and broad confidence intervals.

**Conclusion:** Heat stress is increasing faster than Apparent Temperature across Thailand. While composite measures like the Apparent Temperature and Humidex are stronger predictors of national-level mortality, regional variations in these relationships underscores the need to develop local heat metric that account for both local characteristics and climate change patterns to accurately tailor public health responses.

**Graphical abstract:** 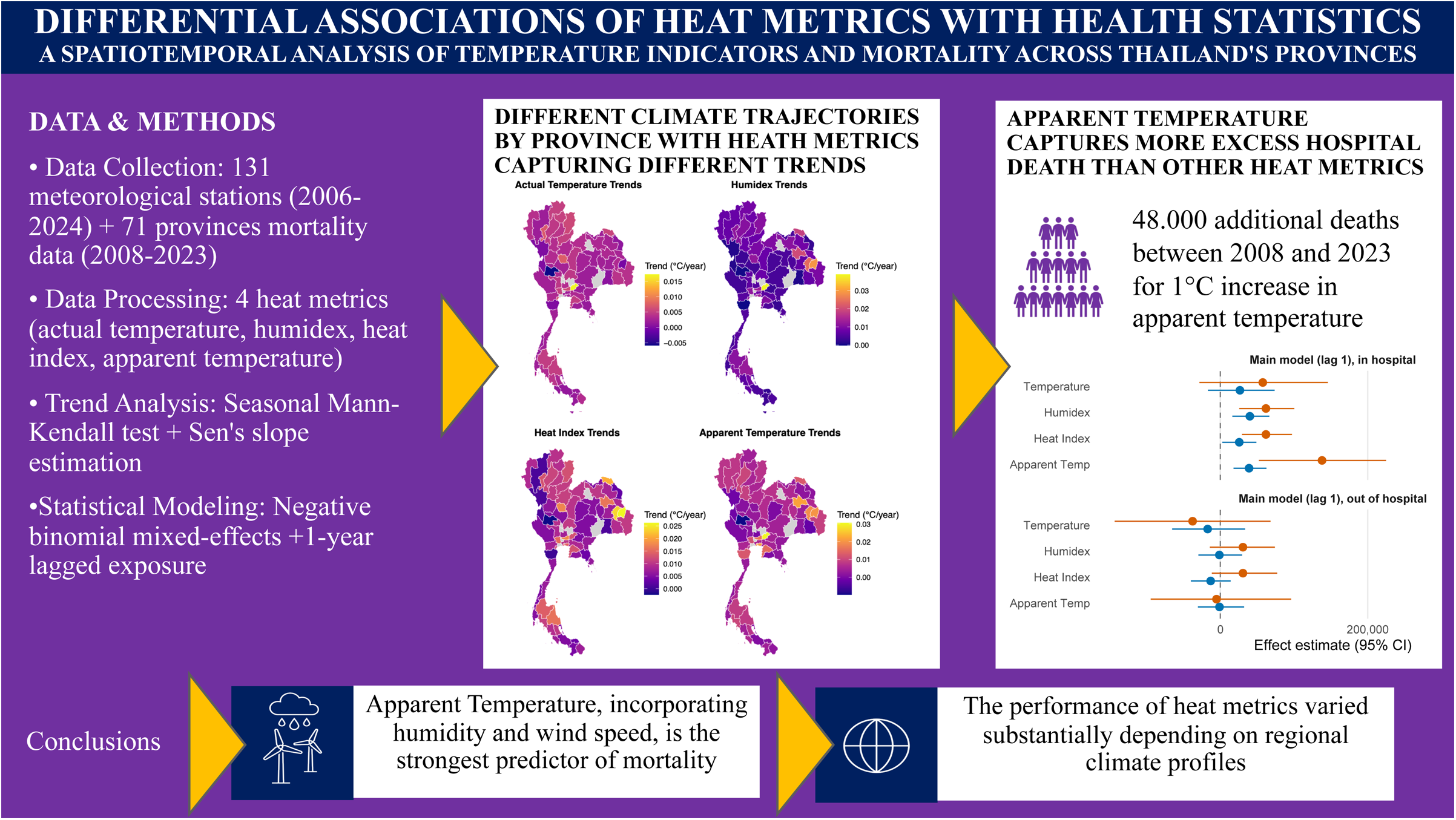

## Background

Southeast Asia is identified as a region highly vulnerable to climate change, with increasing temperatures posing significant threats to population health, economic stability, and environmental systems (Marks, 2011). Within this region, Thailand faces particularly acute challenges. The country has experienced an increase in the frequency and intensity of extreme weather events, including droughts and heatwaves, which are projected to intensify under future climate scenarios (de Oliveira-Júnior et al., 2025; Promchote et al., 2016). The 2011 great flood, for instance, served as a stark indicator of the country’s exposure to climate-induced disasters (Promchote et al., 2016). Recent analyses confirm significant temperature increasing trends across Thailand’s six regions (Kliengchuay et al., 2024). This warming is not uniform, and is exacerbated in urban areas like the Bangkok Metropolitan Region, where the Urban Heat Island (UHI) effect elevates local temperatures due to land-use and land-cover changes (Arifwidodo and Chandrasiri, 2020; Khamchiangta and Dhakal, 2020; Thanvisitthapon et al., 2023). The combined effect of background climate change and urban warming creates a potent threat to the health of the Thai population (Arifwidodo and Chandrasiri, 2020).

Ambient air temperature rising is a crucial metric; however, it presents an incomplete picture of human heat exposure and associated health risks. Humidity strongly influences the human body’s thermoregulatory capacity because high atmospheric moisture levels reduces the evaporation of sweat, the primary mechanism for cooling (Brooke Anderson et al., 2013). Consequently, composite indices that integrate temperature and humidity, such as the Heat Index (HI), provide a more physiologically relevant measure of Apparent Temperature or perceived heat (Brooke Anderson et al., 2013; Lanzante, 2024; Lu and Romps, 2022; Steadman, 1984).

The impacts of this varying exposure on health are severe and well-documented. Elevated temperatures and a high diurnal temperature range are directly linked to increased hospital admissions in Bangkok (Phosri et al., 2020a, 2020b). Most critically, a large national cohort study of Thai workers found a definitive association between heat stress and an increased risk of occupational injury, demonstrating the tangible economic and health consequences of heat exposure (Tawatsupa et al., 2013). The recent finding of a temporal relationship between ambient temperature and non-accidental mortality in Thailand provides further evidence of inadequate adaptation to increasing heat (Sritong-aon et al., 2025). These results are comparable in other counties, looking, for instance; at all causes of mortality across children under 5 years old in six climate African regions (Brimicombe et al., 2024b), London (England) (Simpson et al., 2025), Russia (Timonin et al., 2025) or Europe (Masselot et al., 2023).

The scientific foundation for calculating the Heat Index relies on approximations of saturation vapor pressure, which is itself temperature dependant (Alduchov and Eskridge, 1996; Buck, 1981). These calculations are essential for converting meteorological data into a metric that accurately reflects human experience. Globally, studies have demonstrated that trends in Apparent Temperature can diverge significantly from trends in air temperature alone, with Apparent Temperature often increasing at a faster rate due to concomitant rises in humidity (Jaswal et al., 2017; Li et al., 2018). This has been observed in regional contexts similar to Thailand, such as Vietnam, where Heat Index projections show a more dramatic increase compared to temperature projections, highlighting the compounding effect of humidity (Hoang et al., 2022). Furthermore, different methods for calculating the Heat Index can yield varying results, and the choice of index itself can impact public health warnings, indicating a need for careful selection of metrics (Brooke Anderson et al., 2013; Burgstall et al., 2019; Lanzante, 2024).

Thailand exhibits significant climatic diversity across its provinces, driven by differences in geography, latitude, and urbanization, which results on varying exposures to heat stress. Analyses reveal distinct trends in temperature, humidity, and precipitation across the country’s six regions, confirming that the experience of climate change is not nationally uniform (Kliengchuay et al., 2024; Phumkokrux and Trivej, 2024). For example, the north and northeast face threats of increasing drought (Moazzam et al., 2020; Raksapatcharawong et al., 2020), while urban centres like Bangkok face intensifying heat stress.

Despite the established evidence, critical gaps in the literature necessitate a provincial-level comparison of temperature and Heat Index in Thailand. First, while previous studies have analysed temperature or humidity trends (Kliengchuay et al., 2024), there is a lack of a unified analysis that directly compares the spatial and temporal trends of raw temperature against the composite Heat Index across all provinces. This comparison is essential for identifying regions where the perceived heat risk is escalating faster than what temperature data alone would suggest. Second, the health impact studies conducted in Thailand have often relied on single metrics – either temperature (Sritong-aon et al., 2025) or a composite index (Arifwidodo and Chandrasiri, 2020) without directly comparing the explanatory power of different indicators for health outcomes across different provincial contexts.

Similarly, important gaps exist when looking at the international literature as actual heat remain the preferred indicator(Brimicombe et al., 2024a; Cole et al., 2025; Goddard et al., 2023; Masselot et al., 2023; Narayanan and Keellings, 2025; Simpson et al., 2025; Xiang et al., 2014) and including humidity information is recognised to bring added value (Burgstall et al., 2019). Determining which metric is a better predictor of adverse health outcomes in different regions (e.g., humid coastal provinces vs. drier inland provinces) is crucial for developing targeted public health interventions and early warning systems (Bunting et al., 2024). To address this gap, this study has three primary research questions:

R.Q.1. Do historical trends in ambient air temperature and other heat indicators (Humidex, Heat Index and Apparent Temperatures) vary across all provinces of Thailand and is the province-level magnitude of time change trends uniform depending on the indicators used?
R.Q.2. Are temporal associations between ambient temperature, Apparent Temperature, the Heat Index, Humidex and mortality similar over a 15 year window across Thailand when accounting for provinces’ baseline characteristics?
R.Q.3. Do provinces with different heat change trajectories have different vulnerability to temperature changes?

## Data & Methods

### Data

We used daily gridded meteorological data spanning 1 January 2006 to 14 November 2024, derived from observations collected at 131 weather stations across Thailand (<u>Supplementary</u> <u>File S.1</u>). Each station was georeferenced and assigned to its corresponding Thai province to generate province-level meteorological indicators. Due to missing data at some weather stations, the study covers 71 out of 77 provinces. The dataset includes daily measurements of average temperature (°C), relative humidity (%), and wind speed (m/s). These variables were used to construct four heat indicators: actual Temperature, Humidex, Heat Index, and Apparent Temperature.

Annual all-cause mortality counts within and outside hospitals, and mid-year population estimates for each province between 2008 and 2023 were obtained from official reports of the Thai Ministry of Public Health (Ministry of Public Health, 2024, 2023, 2018, 2013).

### Heat indicators

In this study, we compared four measures of heat commonly used to describe human-perceived thermal stress:

– **Actual temperature** (T) represents the ambient air temperature alone.
– **Humidex**, developed in Canada, combines air temperature and humidity to quantify perceived warmth, using dew point as a measure of atmospheric moisture (Masterton and Richardson CLI, 1979).
– **Heat Index** (HI), developed by Steadman and adopted by the U.S. National Weather Service, also combines temperature and humidity but relies on relative humidity. It was designed to approximate the body’s reduced cooling efficiency under shaded, low-wind conditions (Brooke Anderson et al., 2013; Hoang et al., 2022; Lu and Romps, 2022).
– **Apparent Temperature** (AT) extends these measures by incorporating wind speed alongside temperature and humidity, providing a more comprehensive estimate of thermal comfort under real-world conditions, as wind can enhance heat loss and modify perceived temperature (Jacobs et al., 2013; Sivapragasam and Natarajan, 2021; Steadman, 1984).

Further methodological details are provided in Supplementary File S.2.1.

Although all heat metrics are expressed in °C, they differ in their distributions. To make them directly comparable, we standardized each measure into Z-scores (subtracting the mean and dividing by the standard deviation). This rescaling sets each variable to have mean 0 and standard deviation 1, so that regression coefficients represent the association with the outcome per 1 standard deviation increase in the predictor. We report both the natural (per °C) and standardized (per SD) associations as the former is directly interpretable in real-world units, while the latter allows a fair comparison of the relative predictive strength across metrics.

We implemented a quality control procedure to remove physiologically implausible values outside to each indicator’s defined range (< −10°C or > 50°C). Missing data are shown in <u>Supplementary file S.2.2</u>. Data coverage was high across all meteorological variables, with one notable systematic gap: a systematic temperature data gap occurred in 2016, where no provincial temperature records are available for the entire year across all provinces. Outside of 2016, temperature data showed near-complete coverage, with 68 provinces maintaining ≥95% data availability. Only 9 provinces had partial records: Nakhon Nayok (started 2015), Nong Bua Lamphu (started 2013), Uthai Thani (started 2015), Yasothon (started 2019), Amnat Charoen (started 2020), Bueng Kan (started 2020), Samut Songkhram (started 2017), and Krabi and Phang Nga with minor early-year gaps (2006-2007). Despite later starts, all maintained ≥85% coverage during operational periods. Relative humidity and wind speed data demonstrated virtually complete coverage throughout the entire study period (2006-2025), with 75 provinces achieving ≥99% data completeness for both variables. Only Krabi (99.2% humidity) and Phang Nga (99.7% humidity) showed minimal gaps, missing 59 and 2 days respectively over the 20-year period. Samut Prakan (99.8% wind) and Uttaradit (99.97% wind) showed negligible gaps. 70 provinces (91%) maintained complete (100%) records for both humidity and wind speed throughout the entire monitoring period, including 2016. The consistent high-quality coverage across humidity and wind variables, coupled with only one systematic temperature gap, ensures robust geographical representation and statistical reliability for climate trend analysis.

### Mortality

The mortality records systematically distinguish between in-hospital and out-of-hospital deaths, enabling us to assess total mortality as well as stratified outcomes by place of death. This feature allowed us to replicate analyses separately for in-hospital and out-of-hospital deaths as a sensitivity analysis, providing a more comprehensive evaluation of heat-related mortality risk. The data, collected at province-level, contains no missing value except for two provinces: no information about the full population in Yasothon in 2008 (we imputed 2009 values) and no information on population and deaths counts in Buengkan from 2008 to 2010 (we imputed 2011 values).

### Analysis 1: Provincial Heat Trends Comparison

We conducted a comparative analysis of trends across the different heat metrics for each province. The analytical framework for each indicator relied on the Seasonal Mann-Kendall test, a non-parametric method robust to non-normal data and seasonal cycles (Longobardi and Villani, 2010; Petrovic et al., 2016), combined with Sen’s slope estimation to quantify the magnitude of change (Atta-ur-Rahman and Dawood, 2017). For each province and each heat indicator, the monthly time series was analysed to test the null hypothesis of no monotonic trend. Significance was assessed at α = 0.05. The rate of change for each significant trend was quantified using Sen’s slope, reported in units specific to each indicator per year. For each province, we calculated pairwise differences in Sen’s slope to determine if the perceived warming trend differed significantly depending on the metric used. The statistical significance of these differences was assessed through error propagation, approximating p-values using the normal distribution. Results were visualized through comparative plots and maps to illustrate the spatial distribution of trends and to highlight provinces where the choice of heat indicator meaningfully altered the estimated trend magnitude.

### Analysis 2: Modelling Mortality Associations across Heat Indicators

We fitted a series of negative binomial mixed-effects regression models (Lee et al., 2023) to evaluate the association between annual mortality counts and different heat indicators across provinces, specifying the total number of deaths per province-year as the outcome and using a log link function with an offset for the logarithm of the population to account for varying population sizes (Wels and Hamarat, 2025). The models included a single heat metric as the primary predictor, entered as a continuous variable, with separate models for each indicator to allow direct comparisons. Each model incorporated fixed effects for year to adjust for nationwide temporal trends and province-specific random intercepts to account for baseline differences in mortality rates across provinces, as well as random slopes for the heat indicator to permit province-specific deviations in the strength of the association, thereby acknowledging that the same unit increase in a heat metric may have differential effects in cooler versus hotter regions. To assess delayed effects, we fitted additional models with 1-year, 2-year, and 3-year lags of each heat indicator. In this study, we focus on the 1-year lag because it provides more robust estimates and matches the annual mortality reporting data. Marginal effects were calculated as the expected change in total deaths across all provinces and the study period associated with a 1°C / 1 SD increase in the heat indicator. Predicted death counts were generated for two scenarios: the observed annual values, and the same values increased by one unit. All predictions incorporated the population offset. Confidence intervals were derived via parametric bootstrapping. Model performance and comparative fit across indicators and lag structures were evaluated using the Akaike Information Criterion (AIC) to identify the metric and lag structure most strongly associated with population-level mortality.

### Analysis 3: Modelling Mortality Associations by cluster of heat trajectory profiles

Building upon our previous province-level analyses, we examined whether the temperature-mortality relationship varied across regions with distinct climate trajectories. Our approach consisted of two phases: clustering and effect-modification testing. First, we identified regional climate patterns through k-means clustering of the four heat trajectory metrics. After standardizing these variables using Z-score normalization, we determined the optimal cluster solution (k=4) via the elbow method, which revealed distinct patterns of climate change across Thai provinces. Second, we tested for effect modification by incorporating temperature-cluster interaction terms within negative binomial mixed-effects models. The full specification included temperature metrics interacted with cluster membership, while maintaining controls for yearly variations, population size (offset), and province-level baseline differences (random intercepts). Because population counts vary by province, marginal effects were calculated as the yearly prevalence in excess death. This approach allowed us to formally test whether the temperature-mortality relationship differed significantly across regions experiencing distinct climate trajectories, while preserving the hierarchical structure of provinces nested within climate-pattern groups.

### Software

All analyses were implemented in R version 4.3.0. Data processing used ‘dplyr’ and ‘tidyr’. Seasonal Mann-Kendall tests and Sen’s slope estimation used ‘trend’ and ‘zyp’. Regression modelling was performed with ‘lme4’, and marginal effects were calculated using ‘ggeffects’ and bootstrapping with ‘MASS’. Visualization was done with ‘ggplot2’ and ‘sf’ for spatial mapping.

## Results

### Provincial Heat Trends Comparison

Seasonal trend analysis of temperature, Humidex, Heat Index, and Apparent Temperature across Thailand revealed distinct spatial patterns in warming. Heat stress metrics generally increasing more rapidly than mean temperature as can be seen in **figure 1**; the full estimates by heat metrics and differences across metrics are shown in <u>supplementary file S.3</u>. Four clusters of heat trajectories were generated based on these data, as shown in **figure 2** with details on principal components analysis shown in <u>supplementary file S.4</u>.

**Figure 1.**
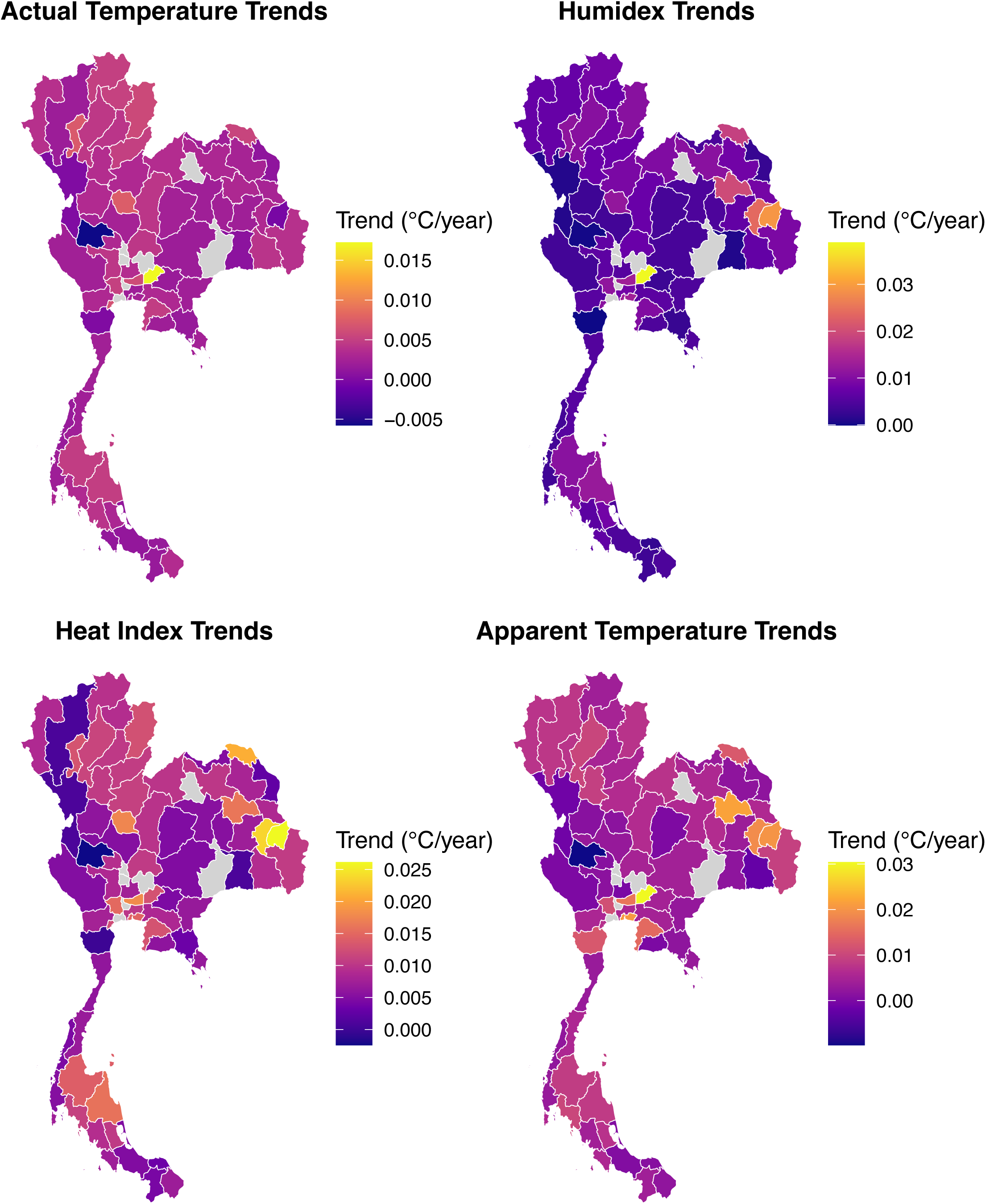
Seasonal Mann-Kendall test by heat indicators by Province.

**Figure 2.**
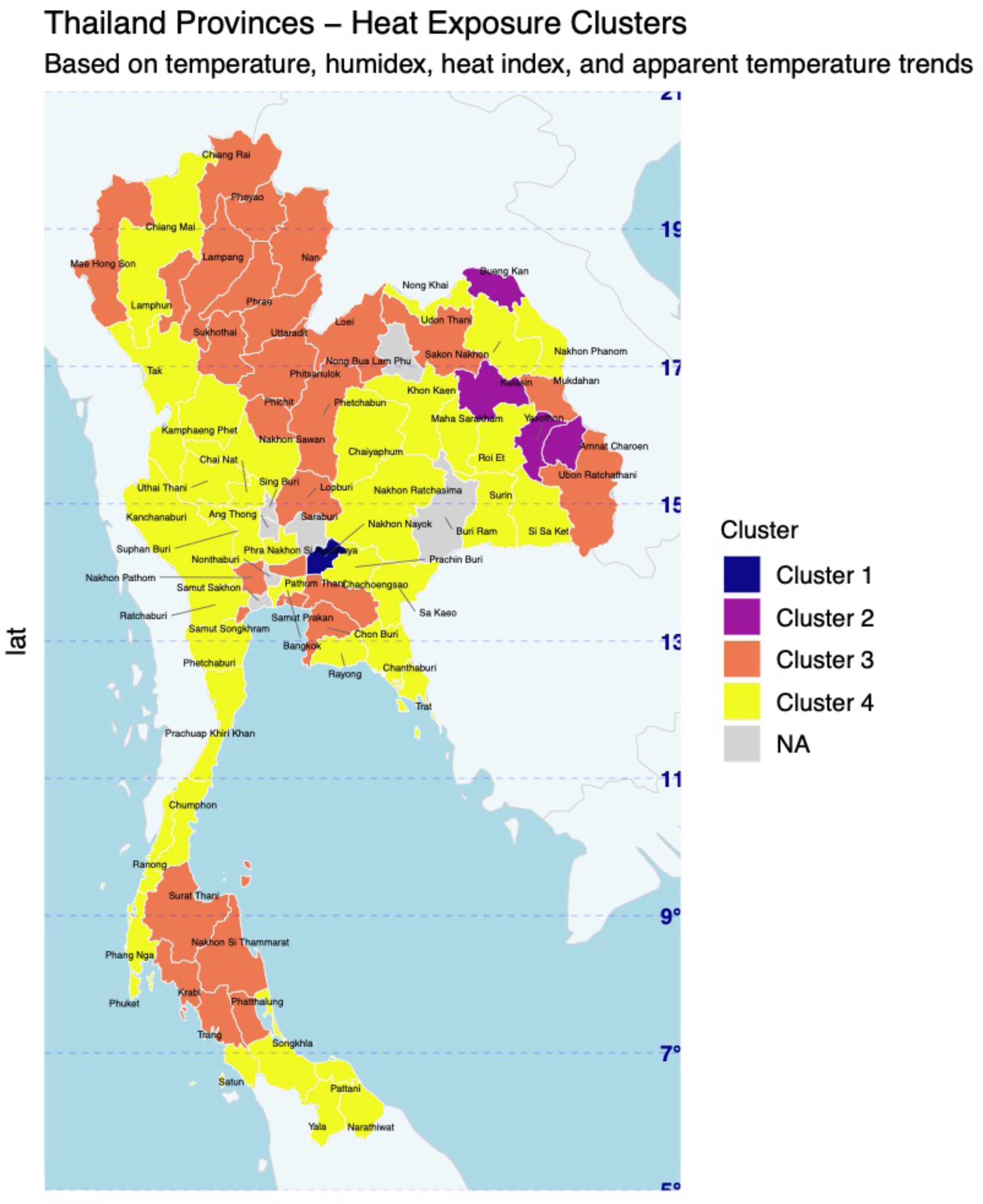
Heat exposures trajectory clusters.

Across the 71 selected provinces, Humidex, Heat Index, and Apparent Temperature trends exceeded mean temperature trends in 63, 67, and 56 provinces, respectively, with differences ranging from 0.001 to 0.038 °C per year. These differences suggest that perceived heat exposure is intensifying more quickly than ambient temperature alone, particularly in northeastern and central regions. For instance, provinces such as Yasothon, Kalasin, and Amnat Charoen exhibited the largest disparities, with Humidex trends exceeding mean temperature trends by 0.027, 0.017, and 0.038 °C per year, respectively.

Significant warming of humid heat metrics was observed in many provinces even when mean temperature trends were weak or non-significant, underscoring the importance of these metrics in capturing nuanced climate signals. In Amnat Charoen, for example, the mean temperature trend was not statistically significant from zero (−0.000 °C per year), while the Humidex increased at a much faster and significant rate of 0.038 °C per year (95% CI 0.033 to 0.043). In Kalasin, a modest mean temperature trend (0.003 °C per year) contrasted with a significant Humidex trend of 0.020 °C per year (95% CI 0.017 to 0.023). Conversely, in provinces such as Uthai Thani, Si Sa Ket, and Phichit, Apparent Temperature trends were negative relative to mean temperature, reflecting a small subset of regions where seasonal factors or local microclimates may counteract general warming trends. These exceptions highlight the heterogeneous nature of seasonal climate change across Thailand.

Spatially, the warming pattern exhibited clear regional gradients. The northeastern and central provinces displayed the most pronounced accelerations in Humidex and Heat Index, creating a pattern of accelerated heat stress in these densely populated and agricultural regions. Northern provinces generally displayed more moderate, though still significant, warming across all metrics. Southern coastal provinces, by contrast, showed smaller differences between mean temperature and heat metrics, with some areas such as Narathiwat and Songkhla exhibiting only minor increases or slight reductions in Apparent Temperature trends relative to mean temperature.

Several provinces in the northeast and central plains, including Nong Bua Lamphu, Loei, and Samut Prakan, demonstrated Humidex and Heat Index trends consistently exceeding mean temperature trends by 0.009 to 0.014 °C per year. Conversely, in northern highland provinces such as Chiang Mai and Nan, differences, while positive, were typically more modest at 0.004–0.005 °C per year, indicating consistent but less accelerated warming in perceived temperature. In southern provinces like Surat Thani and Krabi, Heat Index trends were significantly higher than mean temperature, generally in the 0.008–0.010 °C per year range (95% CI 0.007 to 0.012), while in some provinces like Tak, Apparent Temperature trends were effectively unchanged.

### Mortality Associations across Heat Indicators

The main analyses of 1-year lagged associations are shown in supplementary file, with full results on the natural and Z-score metrics shown in supplementary files S.6 and S.7.

**Figure 3.**
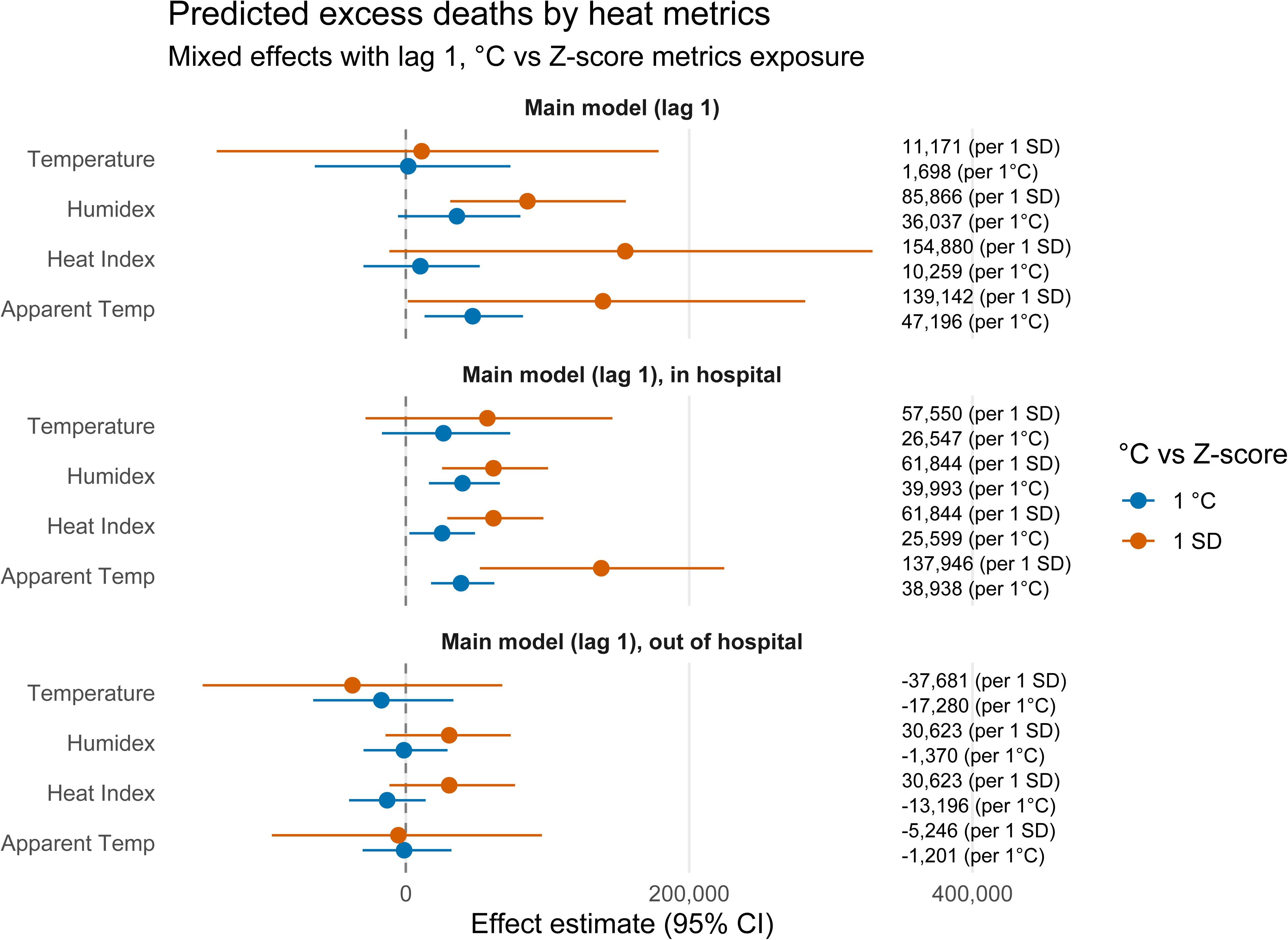
Predicated total excess deaths (ED), hospital ED and non-hospital ED by heat metrics, C° versus Z-score (Mixed effects model with 1-year lag)

Looking at Z-scores transformed exposures, we observe that the Heat Index was associated with an estimated 154,880 Excess Deaths (ED) (95% CI: −11,616 to 329,428) over the study period, followed by Apparent Temperature (139,142 ED; 95% CI: 1,409 to 281,896) and Humidex (85,866 ED; 95% CI: 31,334 to 155,285). The association for mean temperature was weak and non-statistically significant (11,171; 95% CI: −133,462 to 178,431). Analysis using natural-scale metrics, representing a 1°C increase, showed a similar pattern, with Apparent Temperature showing the strongest and only statistically significant association (47,196 ED; 95% CI: 13,287 to 82,704). The point estimates for Humidex (36,037 ED; 95% CI: −5,458 to 80,859) and Heat Index (10,259 ED; 95% CI: −29,902 to 52,110) were positive but non-statistically significant, while the mean temperature estimate was negligible (1,698 ED; 95% CI: −64,217 to 73,797).

This association was overwhelmingly driven by in-hospital mortality. For in-hospital deaths using natural-scale metrics, both Humidex (39,993 ED; 95% CI: 16,302 to 66,404) and Apparent Temperature (38,938 ED; 95% CI: 17,852 to 62,495) showed significant positive associations per 1°C increase. The Z-score models for in-hospital deaths were even more pronounced, with Apparent Temperature associated with 137,946 Excess Deaths (95% CI: 52,289 to 224,764) per 1 SD increase. In contrast, for out-of-hospital deaths, no metrics showed a significant association. All confidence intervals for out-of-hospital deaths straddled the null value. A comparison of model fit statistics confirmed that models incorporating humidity-based metrics consistently outperformed those using mean temperature alone (<u>supplementary file S.8</u>).

### Mortality Associations by cluster of heat trajectory profiles

Cluster-specific analyses revealed that the predictive performance of heat metrics was not uniform across Thailand and was modified by the underlying regional heat trajectory profile. In Cluster 2, which includes provinces with pronounced accelerations in humid heat, mean temperature was the strongest predictor in Z-score models, with an estimate of 0.533 Excess Deaths per 1,000 population (ED per 1,000) (95% CI: −0.300 to 1.737). Humidex also performed well (0.255 ED per 1,000; 95% CI: 0.035 to 0.613). Conversely, in Cluster 3, composite metrics were clearly superior. The Z-score models for Humidex and Heat Index showed significant associations (0.148 ED per 1,000 for both; 95% CI: 0.075 to 0.260 and 0.062 to 0.270, respectively), while the mean temperature estimate was negative and non-significant (−0.063 ED per 1,000; 95% CI: −0.257 to 0.144). Cluster 1 showed positive point estimates for all metrics in Z-score models, with the strongest for Apparent Temperature (0.230 ED per 1,000; 95% CI: −0.033 to 0.657). Cluster 4 exhibited more modest associations, with Apparent Temperature showing the strongest point estimate (0.170 ED per 1,000; 95% CI: −0.026 to 0.401).

**Figure 4.**
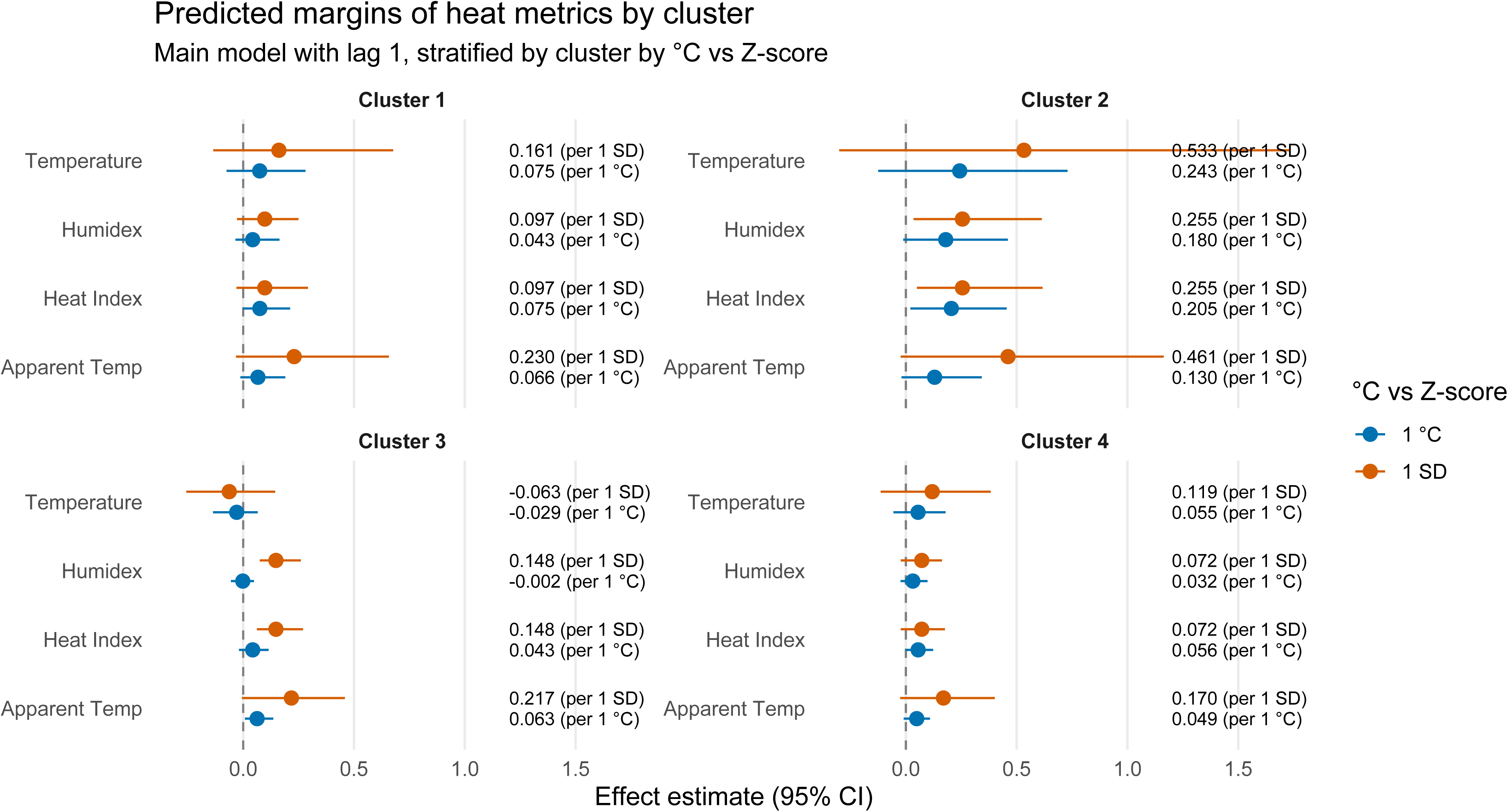
Predicted total death margins by heat metrics and heat change clusters, C° and Z-scores (Mixed effects model with 1-year lag)

This heterogeneity was further emphasized and clarified in models for in-hospital deaths as can be seen in the <u>supplementary files S.6 and S.7</u>. For instance, in Cluster 3, the Z-score model for Apparent Temperature was a strong and significant predictor of in-hospital mortality (0.230 ED per 1,000; 95% CI: 0.123 to 0.399), vastly outperforming mean temperature (0.085 ED per 1,000; 95% CI: −0.047 to 0.230). Similarly, in Cluster 2, the natural-scale model for in-hospital deaths showed a significant association with mean temperature (0.145 ED per 1,000; 95% CI: 0.004 to 0.486). Conversely, analyses of out-of-hospital mortality consistently failed to show significant associations across all clusters and metrics.

## Discussion

This study provides a provincial-level analysis of heat trends and their association with mortality in Thailand, addressing critical gaps in the literature by directly comparing ambient temperature with composite heat metrics including humidity and wind speed. Our findings reveal three insights that challenge the use of a one-size-fits-all metric for heat-health early warning systems and underscore the nuanced nature of climate change impacts across the country.

First, our trend analysis (R.Q.1) confirms that the choice of heat metric fundamentally alters the perceived magnitude and spatial pattern of warming. The consistent finding that Humidex, Heat Index, and Apparent Temperature trends outpace raw temperature trends in most provinces demonstrates that the physiological experience of heat stress is intensifying faster than thermometers alone would suggest. This is particularly acute in the agricultural northeastern and central regions, where humidity appears to be a significant compounding factor. This spatial heterogeneity means that vulnerability assessments based solely on temperature data likely underestimate the true risk in many provinces and overestimate it in others.

Second, mortality analysis (R.Q.2) identified one-year lagged Apparent Temperature as the strongest predictor of all-cause mortality at the national level. The estimated of 47,196 Excess Deaths (95% CI: 13,287 to 82,704) associated with a 1°C increase in this metric underscores the severe population health burden of heat stress in Thailand. This finding aligns with physiological principles, as Apparent Temperature integrates temperature, humidity, and wind speed and therefore offers a more complete measure of perceived heat stress. This association was overwhelmingly driven by in-hospital deaths, where both Humidex and Apparent Temperature showed significant positive associations per 1°C increase.

However, our third finding (R.Q.3) reveals that this national-level conclusion does not hold uniformly across all provinces. Cluster-specific analyses demonstrate fundamental heterogeneity in the drivers of heat-related mortality. These findings indicate that the optimal predictive metric is contingent on local climate trajectories and that the primary drivers of heat-related mortality—whether dry or humid heat—vary by region.

A key limitation of this study is its reliance on all-cause mortality data, which precludes us from examining cause-specific deaths that are more directly linked to heat exposure. Furthermore, while our models controlled for national yearly trends and time-invariant provincial characteristics, the ecological study design cannot account for all potential province-level time-varying confounders, such as changes in age structure, socioeconomic status, or air pollution. Our analysis employed methods to address confounding: national-year fixed effects control for factors affecting all provinces simultaneously (e.g., a national policy change or the COVID-19 pandemic years), and province-level random intercepts account for all time-invariant differences across provinces such as regional demographic structure, baseline infrastructure or socio-economic inequalities. The province-level random slopes further allow the relationship between heat and mortality to vary, capturing differing regional vulnerabilities. While our model’s structure mitigates many sources of bias, we cannot definitively rule out the potential for residual confounding from these unmeasured, time-varying factors. Finally, yearly mortality statistics do not allow us to address the documented relationship between climate change and seasonality of mortality (Madaniyazi et al., 2024), particularly distinguishing the dry season from the monsoon.

Nevertheless, our results present a nuanced, dual prescription for public health policy and future research. While many countries with more homogenous climates may effectively employ a single national heat metric, Thailand’s significant climatic diversity—from humid coastal regions to drier inland provinces—necessitates a more stratified approach. For national-level assessments and policy planning, Apparent Temperature emerges as the most robust single metric to capture the aggregate health burden of heat across the country’s varied regions. However, for regional and local health interventions and early warning systems, a tailored approach is essential, as the optimal predictive metric varies significantly by cluster. This demonstrates that in geographically and climatically diverse nations—a common feature in Southeast Asia and many parts of the world—the drivers of heat-related mortality are context-specific. Therefore, we recommend that other countries facing similar variability conduct sub-national analyses to identify regionally appropriate metrics. This regional strategy will be crucial for Thailand and other diverse nations to effectively adapt to the escalating threat of heat stress and mitigate its impacts on population health.

## Data Availability

All data produced are available online at:
- The Thai Ministry of Public Health
- Climate data were combined by Dr Sittichai Choosumrong. Data are available upon request.

## Acknowledgments

This work was partially supported by the Reinventing University Program 2024, The Ministry of Higher Education, Science, Research and Innovation (MHESI), Thailand (Grant number R2567A145). The authors would also like to thank the Thai Meteorological Department (TMD) for providing weather. Jacques Wels acknowledges funding from the FNRS (Belgium).

## Supplementary files

Supplementary file S.1 Weather Stations across Thailand Provinces
Supplementary file S.2 Heat indicators and missing data

Supplementary file S.2.1. Heat indicators
Supplementary file S.2.1. Missing data
Supplementary file S.3 Seasonal Mann-Kendall test by heat indicators and differences relative to actual temperatures by Province
Supplementary file S.4. Principal component analyses (clusters)
Supplementary file S.5 Metrics summary across all provinces and years

Supplementary file S.5.1 Minimum, maximum, mean, median, Standard Deviation (SD)
Supplementary file S.5.2 Density plot Supplementary file S.5.3 Distribution plot
Supplementary file S.6. Mixed effect using original heat metrics units

Supplementary file S.6.1. Marginal effect of total excess death explained heat metrics
Supplementary file S.6.2. Marginal effect of excess death per 1,000 people explained heat metrics
Supplementary file S.6.3 Marginal effect of heat on Mortality (total excess deaths across all provinces and years and prevalence per 1000) – restricted to hospital deaths
Supplementary file S.6.4 Marginal effect of excess death per 1,000 people explained heat metrics – restricted to hospital deaths
Supplementary file S.6.5. Marginal effect of heat on Mortality (total excess deaths across all provinces and years and prevalence per 1000) – restricted to out of hospital deaths
Supplementary file S.6.6. Marginal effect of excess death per 1,000 people explained heat metrics – restricted to out of hospital deaths
Supplementary file S.7. Mixed effect using Z-scores transformed heat metrics

Supplementary file S.7.1. Marginal effect of total excess death explained heat metrics
Supplementary file S.7.2. Marginal effect of excess death per 1,000 people explained heat metrics
Supplementary file S.7.3 Marginal effect of heat on Mortality (total excess deaths across all provinces and years and prevalence per 1000) – restricted to hospital deaths
Supplementary file S.7.4 Marginal effect of excess death per 1,000 people explained heat metrics – restricted to hospital deaths
Supplementary file S.7.5. Marginal effect of heat on Mortality (total excess deaths across all provinces and years and prevalence per 1000) – restricted to out of hospital deaths
Supplementary file S.7.6. Marginal effect of excess death per 1,000 people explained heat metrics – restricted to out of hospital deaths
Supplementary file S.8. Main models specification

## Authors statement

Conceptualization: JW, RK

Data Curation: SC, JW

Formal Analysis: JW

Funding Acquisition: RK, JW

Investigation: JW, RK

Methodology: JW, RK

Project Administration: JW, RK, SC

Resources: NA

Software: JW

Supervision: JW

Validation: JGH

Visualization: JW

Writing – Original Draft Preparation: JW

Writing – Review & Editing : RK, JGH

Note: Jacques Wels: JW; Romnalin Keanjoom: RK; Juan Gonzalez Hijon: JGH; Sittichai Choosumrong: SC

