## Supplementary file for "Differential Associations of Heat Metrics with Health Statistics: A Spatiotemporal Analysis of Temperature Indicators and Mortality Across Thailand Provinces"

**SUPPLEMENTARY FILES**

### Supplementary file S.1 Weather Stations across Thailand Provinces

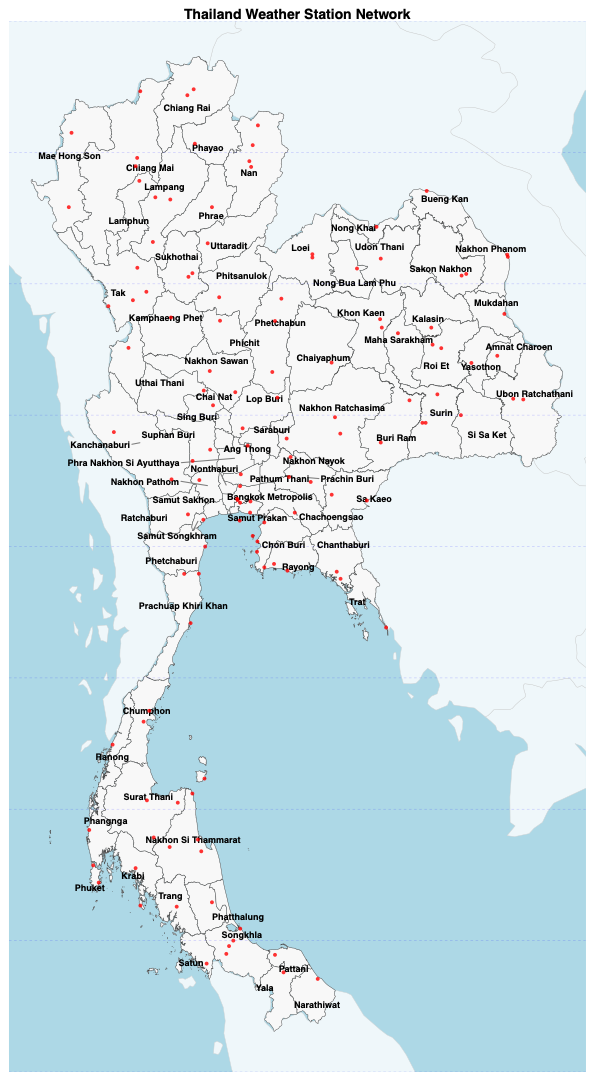

### Supplementary file S.2 Heat indicators and missing data

#### Supplementary file S.2.1. Heat indicators

**Humidex** (short for "humidity index") is a metric developed by Canadian meteorologists to quantify the perceived temperature by humans, accounting for the combined effects of ambient temperature and humidity. Unlike relative humidity, the Humidex uses dew point temperature to provide an absolute measure of atmospheric moisture content, making it a more consistent indicator of mugginess throughout the day. The Humidex is calculated as follows [41]:

Humidex = T + h, where h = (0.5555) × (e - 10.0) and e = 6.112 × 10^(7.5 × Td / (237.7 + Td))

Here, T is the air temperature (°C), Td is the dew point temperature (°C), and e is the vapour pressure in hectopascals (hPa). The dew point (Td) is itself derived from air temperature and relative humidity using the Magnus formula [12,13]. The resulting Humidex value is a dimensionless number interpreted as an "equivalent feel-like" temperature in degrees Celsius.

The **Heat Index** (HI), also known as the "apparent temperature" in some contexts, is a measure developed by Steadman (1979) and adopted by the U.S. National Weather Service (NWS) that represents the human-perceived equivalent temperature based on the combined effects of ambient temperature and relative humidity. It is derived from a physiological model of human heat balance and approximates the body's inability to cool itself effectively through sweat evaporation when humidity is high. The Heat Index is calculated using a complex polynomial regression formula:

HI = c₁ + c₂T + c₃RH + c₄T*RH + c₅T² + c₆RH² + c₇T²*RH + c₈T*RH² + c₉T²*RH²

where T is the air temperature in degrees Fahrenheit (°F), RH is the relative humidity (%), and c₁–c₉ are empirically derived coefficients (c₁ = -42.379, c₂ = 2.04901523, c₃ = 10.14333127, c₄ = -0.22475541, c₅ = -0.00683783, c₆ = -0.05481717, c₇ = 0.00122874, c₈ = 0.00085282, c₉ = -0.00000199). The result is converted back to degrees Celsius for interpretation. The Heat Index is designed for use in shaded, light-wind conditions and provides critical thresholds for heat-related health risks [8].

The **Apparent temperature** (AT)**,** represents the perceived air temperature by humans, accounting for the combined effects of ambient temperature, relative humidity, and wind speed. While traditional heat indices rely solely on temperature and humidity, Steadman’s extended formula incorporates wind speed to provide a more realistic estimate of thermal comfort in warm conditions[9]. The apparent temperature (AT) in degrees Celsius (°C) is calculated as follow:

AT = T + (0.33 × e) - (0.70 × WS) - 4.00

where T is the air temperature (°C), WS is the wind speed (m/s), and e is the water vapor pressure in hPa. The vapor pressure e is derived from relative humidity (RH) (in %) and temperature using the modified Magnus formula:

e = (RH / 100) × 6.105 × exp((17.27 × T) / (237.7 + T))

The coefficients **17.27** and **237.7** in the Magnus formula are empirically derived and validated for temperatures between 0–50°C [12,13]. This formulation reflects how AT increases with humidity (via vapor pressure) and decreases with wind speed, capturing the dual role of moisture and ventilation in perceived heat. This formulation allows apparent temperature to increase with humidity (via vapor pressure) and decrease with higher wind speeds, reflecting the dual influence of moisture and ventilation on perceived heat. Although daily rainfall does not enter the equation directly, it often affects apparent temperature indirectly by increasing near-surface humidity and altering wind patterns following precipitation events. For analytical purposes, rainfall may be included as a contextual variable rather than as an input in the heat index formula itself.

#### Supplementary file S.2.1. Missing data

|  | Temperature | | |  | Humidity | | |  | Wind speed | | |
| --- | --- | --- | --- | --- | --- | --- | --- | --- | --- | --- | --- |
| Province | total days | missing days | missing (pct) |  | total days | missing days | missing (pct) |  | total days | missing days | missing (pct) |
| Amnat Charoen | 1779 | 0 | 0 |  | 1982 | 0 | 0 |  | 2023 | 0 | 0 |
| Bangkok | 6524 | 0 | 0 |  | 7095 | 0 | 0 |  | 7136 | 0 | 0 |
| Bueng Kan | 1738 | 0 | 0 |  | 1954 | 0 | 0 |  | 1995 | 0 | 0 |
| Buriram | 6522 | 0 | 0 |  | 7095 | 0 | 0 |  | 7136 | 0 | 0 |
| Chachoengsao | 6150 | 0 | 0 |  | 7090 | 0 | 0 |  | 7131 | 0 | 0 |
| Chai Nat | 6155 | 0 | 0 |  | 7095 | 0 | 0 |  | 7136 | 0 | 0 |
| Chaiyaphum | 6483 | 0 | 0 |  | 7095 | 0 | 0 |  | 7136 | 0 | 0 |
| Chanthaburi | 6507 | 0 | 0 |  | 7095 | 0 | 0 |  | 7136 | 0 | 0 |
| Chiang Mai | 6521 | 0 | 0 |  | 7095 | 0 | 0 |  | 7136 | 0 | 0 |
| Chiang Rai | 6519 | 0 | 0 |  | 7095 | 0 | 0 |  | 7136 | 0 | 0 |
| Chon Buri | 6524 | 0 | 0 |  | 7095 | 0 | 0 |  | 7136 | 0 | 0 |
| Chumphon | 6520 | 0 | 0 |  | 7095 | 0 | 0 |  | 7136 | 0 | 0 |
| Kalasin | 6466 | 0 | 0 |  | 7066 | 0 | 0 |  | 7107 | 0 | 0 |
| Kamphaeng Phet | 6519 | 0 | 0 |  | 7095 | 0 | 0 |  | 7136 | 0 | 0 |
| Kanchanaburi | 6522 | 0 | 0 |  | 7095 | 0 | 0 |  | 7136 | 0 | 0 |
| Khon Kaen | 6522 | 0 | 0 |  | 7095 | 0 | 0 |  | 7136 | 0 | 0 |
| Krabi | 6419 | 0 | 0 |  | 7079 | 59 | 0.8 |  | 7115 | 0 | 0 |
| Lampang | 6524 | 0 | 0 |  | 7095 | 0 | 0 |  | 7136 | 0 | 0 |
| Lamphun | 6517 | 0 | 0 |  | 7095 | 0 | 0 |  | 7136 | 0 | 0 |
| Loei | 6522 | 0 | 0 |  | 7095 | 0 | 0 |  | 7136 | 0 | 0 |
| Lopburi | 6523 | 0 | 0 |  | 7095 | 0 | 0 |  | 7136 | 0 | 0 |
| Mae Hong Son | 6522 | 0 | 0 |  | 7095 | 0 | 0 |  | 7136 | 0 | 0 |
| Maha Sarakham | 6455 | 0 | 0 |  | 7093 | 0 | 0 |  | 7134 | 0 | 0 |
| Mukdahan | 6508 | 0 | 0 |  | 7095 | 0 | 0 |  | 7136 | 0 | 0 |
| Nakhon Nayok | 3031 | 0 | 0 |  | 2227 | 0 | 0 |  | 2268 | 0 | 0 |
| Nakhon Pathom | 6108 | 0 | 0 |  | 7095 | 0 | 0 |  | 7136 | 0 | 0 |
| Nakhon Phanom | 6524 | 0 | 0 |  | 7095 | 0 | 0 |  | 7136 | 0 | 0 |
| Nakhon Ratchasima | 6524 | 0 | 0 |  | 7095 | 0 | 0 |  | 7136 | 0 | 0 |
| Nakhon Sawan | 6522 | 0 | 0 |  | 7095 | 0 | 0 |  | 7136 | 0 | 0 |
| Nakhon Si Thammarat | 6522 | 0 | 0 |  | 7095 | 0 | 0 |  | 7136 | 0 | 0 |
| Nan | 6524 | 0 | 0 |  | 7095 | 0 | 0 |  | 7136 | 0 | 0 |
| Narathiwat | 5938 | 0 | 0 |  | 7095 | 0 | 0 |  | 7136 | 0 | 0 |
| Nong Bua Lamphu | 3730 | 0 | 0 |  | 5999 | 0 | 0 |  | 6040 | 0 | 0 |
| Nong Khai | 6444 | 0 | 0 |  | 7095 | 0 | 0 |  | 7136 | 0 | 0 |
| Pathum Thani | 6115 | 0 | 0 |  | 7052 | 0 | 0 |  | 7093 | 0 | 0 |
| Pattani | 6510 | 0 | 0 |  | 7095 | 0 | 0 |  | 7136 | 0 | 0 |
| Phang Nga | 6395 | 0 | 0 |  | 7012 | 2 | 0 |  | 7028 | 0 | 0 |
| Phatthalung | 6146 | 0 | 0 |  | 7088 | 0 | 0 |  | 7129 | 0 | 0 |
| Phayao | 6447 | 0 | 0 |  | 7095 | 0 | 0 |  | 7136 | 0 | 0 |
| Phetchabun | 6523 | 0 | 0 |  | 7095 | 0 | 0 |  | 7136 | 0 | 0 |
| Phetchaburi | 6517 | 0 | 0 |  | 7095 | 0 | 0 |  | 7136 | 0 | 0 |
| Phichit | 6138 | 0 | 0 |  | 7095 | 0 | 0 |  | 7136 | 0 | 0 |
| Phitsanulok | 6388 | 0 | 0 |  | 7095 | 0 | 0 |  | 7136 | 0 | 0 |
| Phra Nakhon Si Ayutthaya | 6115 | 0 | 0 |  | 7095 | 0 | 0 |  | 7136 | 0 | 0 |
| Phrae | 6473 | 0 | 0 |  | 7095 | 0 | 0 |  | 7136 | 0 | 0 |
| Phuket | 6524 | 0 | 0 |  | 7095 | 0 | 0 |  | 7136 | 0 | 0 |
| Prachin Buri | 6518 | 0 | 0 |  | 7092 | 0 | 0 |  | 7133 | 0 | 0 |
| Prachuap Khiri Khan | 6517 | 0 | 0 |  | 7095 | 0 | 0 |  | 7136 | 0 | 0 |
| Ranong | 6518 | 0 | 0 |  | 7095 | 0 | 0 |  | 7136 | 0 | 0 |
| Ratchaburi | 6121 | 0 | 0 |  | 7095 | 0 | 0 |  | 7136 | 0 | 0 |
| Rayong | 6524 | 0 | 0 |  | 7095 | 0 | 0 |  | 7136 | 0 | 0 |
| Roi Et | 6523 | 0 | 0 |  | 7095 | 0 | 0 |  | 7136 | 0 | 0 |
| Sa Kaeo | 6521 | 0 | 0 |  | 7095 | 0 | 0 |  | 7136 | 0 | 0 |
| Sakon Nakhon | 6521 | 0 | 0 |  | 7095 | 0 | 0 |  | 7136 | 0 | 0 |
| Samut Prakan | 6508 | 0 | 0 |  | 7095 | 14 | 0.2 |  | 7136 | 0 | 0 |
| Samut Songkhram | 2455 | 0 | 0 |  | 2227 | 0 | 0 |  | 2268 | 0 | 0 |
| Satun | 6455 | 0 | 0 |  | 7095 | 0 | 0 |  | 7136 | 0 | 0 |
| Si Sa Ket | 6134 | 0 | 0 |  | 7089 | 0 | 0 |  | 7130 | 0 | 0 |
| Songkhla | 6527 | 0 | 0 |  | 7095 | 0 | 0 |  | 7136 | 0 | 0 |
| Sukhothai | 6456 | 0 | 0 |  | 7095 | 0 | 0 |  | 7136 | 0 | 0 |
| Suphan Buri | 6520 | 0 | 0 |  | 7095 | 0 | 0 |  | 7136 | 0 | 0 |
| Surat Thani | 6523 | 0 | 0 |  | 7095 | 0 | 0 |  | 7136 | 0 | 0 |
| Surin | 6524 | 0 | 0 |  | 7095 | 0 | 0 |  | 7136 | 0 | 0 |
| Tak | 6524 | 0 | 0 |  | 7095 | 0 | 0 |  | 7136 | 0 | 0 |
| Trang | 6474 | 0 | 0 |  | 7095 | 0 | 0 |  | 7136 | 0 | 0 |
| Trat | 6019 | 0 | 0 |  | 7095 | 0 | 0 |  | 7136 | 0 | 0 |
| Ubon Ratchathani | 6524 | 0 | 0 |  | 7095 | 0 | 0 |  | 7136 | 0 | 0 |
| Udon Thani | 6514 | 0 | 0 |  | 7095 | 0 | 0 |  | 7136 | 0 | 0 |
| Uthai Thani | 2904 | 0 | 0 |  | 2227 | 0 | 0 |  | 2268 | 0 | 0 |
| Uttaradit | 6495 | 0 | 0 |  | 7095 | 2 | 0 |  | 7136 | 0 | 0 |
| Yala | 6107 | 0 | 0 |  | 7095 | 0 | 0 |  | 7136 | 0 | 0 |
| Yasothon | 1774 | 0 | 0 |  | 1983 | 0 | 0 |  | 2024 | 0 | 0 |

### Supplementary file S.3 Seasonal Mann-Kendall test by heat indicators and differences relative to actual temperatures by Province

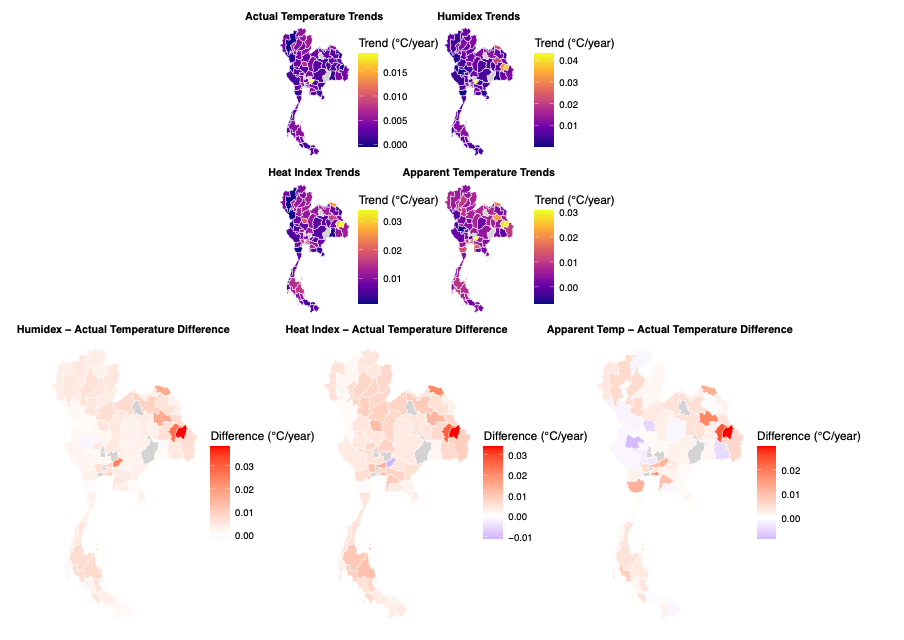

| **Province** | **Actual Temp Trend** | **Humidex Trend** | **Heat Index Trend** | **Apparent Temp Trend** | **Humidex Difference** | **Heat Index Difference** | **Apparent Temp Difference** |
| --- | --- | --- | --- | --- | --- | --- | --- |
| Amnat Charoen | -0.000 °C/yr (τ=-0.006, ns) | 0.038 °C/yr (τ=0.317, *) | 0.034 °C/yr (τ=0.250, ns) | 0.029 °C/yr (τ=0.322, *) | 0.0384532 | 0.0340499 | 0.0296345 |
| Bangkok | 0.002 °C/yr (τ=-0.064, ns) | 0.008 °C/yr (τ=0.035, ns) | 0.010 °C/yr (τ=0.231, *) | 0.006 °C/yr (τ=0.104, *) | 0.0064191 | 0.0078291 | 0.0038775 |
| Bueng Kan | 0.005 °C/yr (τ=0.011, ns) | 0.023 °C/yr (τ=0.172, ns) | 0.026 °C/yr (τ=0.261, ns) | 0.018 °C/yr (τ=0.183, ns) | 0.0172223 | 0.0207561 | 0.0124192 |
| Buriram | 0.003 °C/yr (τ=-0.058, ns) | 0.007 °C/yr (τ=0.200, *) | 0.007 °C/yr (τ=0.199, *) | 0.005 °C/yr (τ=0.165, *) | 0.0038707 | 0.0046864 | 0.0022502 |
| Chachoengsao | 0.003 °C/yr (τ=0.014, ns) | 0.007 °C/yr (τ=0.115, *) | 0.011 °C/yr (τ=0.255, *) | 0.005 °C/yr (τ=0.032, ns) | 0.0041110 | 0.0075764 | 0.0017927 |
| Chai Nat | 0.003 °C/yr (τ=-0.075, ns) | 0.007 °C/yr (τ=0.025, ns) | 0.010 °C/yr (τ=0.241, *) | 0.002 °C/yr (τ=0.012, ns) | 0.0042166 | 0.0074130 | -0.0011638 |
| Chaiyaphum | 0.002 °C/yr (τ=-0.087, ns) | 0.005 °C/yr (τ=0.155, *) | 0.006 °C/yr (τ=0.150, *) | 0.001 °C/yr (τ=0.045, ns) | 0.0028896 | 0.0036277 | -0.0011256 |
| Chanthaburi | 0.001 °C/yr (τ=0.016, ns) | 0.003 °C/yr (τ=-0.021, ns) | 0.003 °C/yr (τ=-0.022, ns) | 0.002 °C/yr (τ=0.020, ns) | 0.0011942 | 0.0021321 | 0.0008928 |
| Chiang Mai | -0.000 °C/yr (τ=-0.047, ns) | 0.004 °C/yr (τ=0.171, *) | 0.001 °C/yr (τ=0.059, ns) | 0.005 °C/yr (τ=0.213, *) | 0.0045973 | 0.0014875 | 0.0056494 |
| Chiang Rai | 0.005 °C/yr (τ=0.329, *) | 0.009 °C/yr (τ=0.316, *) | 0.010 °C/yr (τ=0.342, *) | 0.004 °C/yr (τ=0.147, *) | 0.0036872 | 0.0045847 | -0.0011045 |
| Chon Buri | 0.005 °C/yr (τ=0.153, *) | 0.010 °C/yr (τ=0.105, *) | 0.014 °C/yr (τ=0.364, *) | 0.015 °C/yr (τ=0.303, *) | 0.0048722 | 0.0088152 | 0.0095629 |
| Chumphon | 0.002 °C/yr (τ=0.018, ns) | 0.007 °C/yr (τ=0.044, ns) | 0.008 °C/yr (τ=0.259, *) | 0.006 °C/yr (τ=0.056, ns) | 0.0046790 | 0.0052665 | 0.0040140 |
| Kalasin | 0.003 °C/yr (τ=-0.087, ns) | 0.020 °C/yr (τ=0.239, *) | 0.016 °C/yr (τ=0.165, *) | 0.022 °C/yr (τ=0.261, *) | 0.0166787 | 0.0130289 | 0.0183395 |
| Kamphaeng Phet | 0.003 °C/yr (τ=-0.063, ns) | 0.004 °C/yr (τ=0.027, ns) | 0.008 °C/yr (τ=0.179, *) | 0.002 °C/yr (τ=-0.009, ns) | 0.0008654 | 0.0044986 | -0.0012758 |
| Kanchanaburi | 0.002 °C/yr (τ=-0.006, ns) | 0.004 °C/yr (τ=-0.001, ns) | 0.005 °C/yr (τ=0.144, *) | 0.000 °C/yr (τ=-0.018, ns) | 0.0021260 | 0.0036347 | -0.0012338 |
| Khon Kaen | 0.002 °C/yr (τ=-0.143, *) | 0.006 °C/yr (τ=0.176, *) | 0.007 °C/yr (τ=0.173, *) | 0.007 °C/yr (τ=0.213, *) | 0.0035833 | 0.0044826 | 0.0045519 |
| Krabi | 0.004 °C/yr (τ=0.221, *) | 0.011 °C/yr (τ=0.419, *) | 0.012 °C/yr (τ=0.383, *) | 0.011 °C/yr (τ=0.356, *) | 0.0071101 | 0.0085286 | 0.0073471 |
| Lampang | 0.004 °C/yr (τ=0.164, *) | 0.011 °C/yr (τ=0.377, *) | 0.012 °C/yr (τ=0.330, *) | 0.010 °C/yr (τ=0.433, *) | 0.0061216 | 0.0070610 | 0.0053677 |
| Lamphun | 0.007 °C/yr (τ=0.411, *) | 0.011 °C/yr (τ=0.423, *) | 0.013 °C/yr (τ=0.415, *) | 0.007 °C/yr (τ=0.345, *) | 0.0040903 | 0.0058857 | 0.0002827 |
| Loei | 0.004 °C/yr (τ=-0.022, ns) | 0.011 °C/yr (τ=0.314, *) | 0.011 °C/yr (τ=0.275, *) | 0.008 °C/yr (τ=0.160, *) | 0.0072050 | 0.0076867 | 0.0042774 |
| Lopburi | 0.004 °C/yr (τ=-0.010, ns) | 0.009 °C/yr (τ=0.180, *) | 0.012 °C/yr (τ=0.318, *) | 0.006 °C/yr (τ=0.075, ns) | 0.0045932 | 0.0075936 | 0.0011157 |
| Mae Hong Son | 0.004 °C/yr (τ=0.273, *) | 0.007 °C/yr (τ=0.267, *) | 0.009 °C/yr (τ=0.290, *) | 0.007 °C/yr (τ=0.347, *) | 0.0034110 | 0.0049200 | 0.0036075 |
| Maha Sarakham | 0.002 °C/yr (τ=-0.134, *) | 0.005 °C/yr (τ=-0.021, ns) | 0.007 °C/yr (τ=-0.008, ns) | 0.006 °C/yr (τ=-0.007, ns) | 0.0028258 | 0.0046862 | 0.0046319 |
| Mukdahan | 0.003 °C/yr (τ=0.058, ns) | 0.010 °C/yr (τ=0.257, *) | 0.010 °C/yr (τ=0.230, *) | 0.007 °C/yr (τ=0.211, *) | 0.0069810 | 0.0069452 | 0.0040310 |
| Nakhon Nayok | 0.019 °C/yr (τ=0.139, ns) | 0.043 °C/yr (τ=0.394, *) | 0.008 °C/yr (τ=0.061, ns) | 0.030 °C/yr (τ=0.222, *) | 0.0242754 | -0.0110402 | 0.0117623 |
| Nakhon Pathom | 0.005 °C/yr (τ=0.012, ns) | 0.012 °C/yr (τ=0.074, ns) | 0.016 °C/yr (τ=0.238, *) | 0.011 °C/yr (τ=0.119, *) | 0.0063790 | 0.0104990 | 0.0054661 |
| Nakhon Phanom | 0.001 °C/yr (τ=-0.077, ns) | 0.004 °C/yr (τ=0.049, ns) | 0.004 °C/yr (τ=0.063, ns) | 0.003 °C/yr (τ=0.040, ns) | 0.0028035 | 0.0029081 | 0.0021222 |
| Nakhon Ratchasima | 0.002 °C/yr (τ=-0.072, ns) | 0.005 °C/yr (τ=0.184, *) | 0.006 °C/yr (τ=0.198, *) | 0.004 °C/yr (τ=0.055, ns) | 0.0028802 | 0.0037540 | 0.0022209 |
| Nakhon Sawan | 0.003 °C/yr (τ=-0.117, *) | 0.005 °C/yr (τ=-0.007, ns) | 0.007 °C/yr (τ=0.197, *) | 0.003 °C/yr (τ=-0.001, ns) | 0.0019994 | 0.0045064 | 0.0009503 |
| Nakhon Si Thammarat | 0.005 °C/yr (τ=0.293, *) | 0.012 °C/yr (τ=0.390, *) | 0.016 °C/yr (τ=0.357, *) | 0.008 °C/yr (τ=0.304, *) | 0.0075059 | 0.0109706 | 0.0034479 |
| Nan | 0.006 °C/yr (τ=0.380, *) | 0.011 °C/yr (τ=0.392, *) | 0.013 °C/yr (τ=0.396, *) | 0.007 °C/yr (τ=0.325, *) | 0.0048929 | 0.0069005 | 0.0015417 |
| Narathiwat | 0.004 °C/yr (τ=0.037, ns) | 0.005 °C/yr (τ=0.082, ns) | 0.007 °C/yr (τ=0.058, ns) | 0.002 °C/yr (τ=-0.012, ns) | 0.0011494 | 0.0035596 | -0.0013914 |
| Nong Bua Lamphu | 0.004 °C/yr (τ=0.066, ns) | 0.019 °C/yr (τ=0.340, *) | 0.019 °C/yr (τ=0.286, *) | 0.016 °C/yr (τ=0.256, *) | 0.0144309 | 0.0150824 | 0.0112942 |
| Nong Khai | 0.002 °C/yr (τ=-0.039, ns) | 0.006 °C/yr (τ=0.030, ns) | 0.007 °C/yr (τ=0.027, ns) | 0.005 °C/yr (τ=0.014, ns) | 0.0038875 | 0.0049282 | 0.0025094 |
| Pathum Thani | 0.006 °C/yr (τ=0.072, ns) | 0.014 °C/yr (τ=0.187, *) | 0.020 °C/yr (τ=0.298, *) | 0.016 °C/yr (τ=0.312, *) | 0.0080406 | 0.0140824 | 0.0102849 |
| Pattani | 0.002 °C/yr (τ=0.106, *) | 0.002 °C/yr (τ=-0.084, ns) | 0.004 °C/yr (τ=0.116, *) | 0.001 °C/yr (τ=-0.054, ns) | 0.0006443 | 0.0022357 | -0.0006635 |
| Phang Nga | 0.002 °C/yr (τ=0.115, *) | 0.003 °C/yr (τ=0.168, *) | 0.005 °C/yr (τ=0.110, *) | 0.003 °C/yr (τ=0.117, *) | 0.0011649 | 0.0027647 | 0.0006552 |
| Phatthalung | 0.003 °C/yr (τ=0.218, *) | 0.009 °C/yr (τ=0.352, *) | 0.010 °C/yr (τ=0.282, *) | 0.007 °C/yr (τ=0.335, *) | 0.0056228 | 0.0071974 | 0.0040665 |
| Phayao | 0.005 °C/yr (τ=0.005, ns) | 0.008 °C/yr (τ=0.152, *) | 0.009 °C/yr (τ=0.086, ns) | 0.005 °C/yr (τ=0.108, *) | 0.0027288 | 0.0041456 | -0.0002294 |
| Phetchabun | 0.004 °C/yr (τ=-0.049, ns) | 0.009 °C/yr (τ=0.295, *) | 0.011 °C/yr (τ=0.312, *) | 0.007 °C/yr (τ=0.071, ns) | 0.0047013 | 0.0069586 | 0.0024871 |
| Phetchaburi | 0.001 °C/yr (τ=-0.065, ns) | 0.000 °C/yr (τ=-0.039, ns) | 0.001 °C/yr (τ=0.031, ns) | 0.013 °C/yr (τ=0.290, *) | -0.0005701 | 0.0006449 | 0.0120813 |
| Phichit | 0.007 °C/yr (τ=0.057, ns) | 0.012 °C/yr (τ=0.282, *) | 0.019 °C/yr (τ=0.400, *) | 0.002 °C/yr (τ=0.046, ns) | 0.0050274 | 0.0116119 | -0.0051706 |
| Phitsanulok | 0.003 °C/yr (τ=-0.007, ns) | 0.009 °C/yr (τ=0.078, ns) | 0.011 °C/yr (τ=0.028, ns) | 0.006 °C/yr (τ=0.129, *) | 0.0064727 | 0.0081907 | 0.0033914 |
| Phra Nakhon Si Ayutthaya | 0.004 °C/yr (τ=0.035, ns) | 0.005 °C/yr (τ=-0.029, ns) | 0.009 °C/yr (τ=0.274, *) | 0.000 °C/yr (τ=-0.060, ns) | 0.0018561 | 0.0053679 | -0.0030184 |
| Phrae | 0.004 °C/yr (τ=0.042, ns) | 0.009 °C/yr (τ=0.306, *) | 0.011 °C/yr (τ=0.308, *) | 0.006 °C/yr (τ=0.220, *) | 0.0050231 | 0.0070429 | 0.0013464 |
| Phuket | 0.003 °C/yr (τ=0.209, *) | 0.006 °C/yr (τ=0.192, *) | 0.008 °C/yr (τ=0.243, *) | 0.006 °C/yr (τ=0.210, *) | 0.0029010 | 0.0050273 | 0.0034300 |
| Prachin Buri | 0.002 °C/yr (τ=-0.089, ns) | 0.004 °C/yr (τ=0.003, ns) | 0.006 °C/yr (τ=0.202, *) | 0.005 °C/yr (τ=0.031, ns) | 0.0021319 | 0.0045557 | 0.0029939 |
| Prachuap Khiri Khan | 0.002 °C/yr (τ=0.018, ns) | 0.005 °C/yr (τ=-0.008, ns) | 0.007 °C/yr (τ=0.252, *) | 0.003 °C/yr (τ=0.031, ns) | 0.0027631 | 0.0043890 | 0.0006465 |
| Ranong | 0.002 °C/yr (τ=0.088, ns) | 0.005 °C/yr (τ=0.181, *) | 0.006 °C/yr (τ=0.106, *) | 0.003 °C/yr (τ=0.044, ns) | 0.0034055 | 0.0042072 | 0.0013740 |
| Ratchaburi | 0.003 °C/yr (τ=0.035, ns) | 0.006 °C/yr (τ=0.046, ns) | 0.009 °C/yr (τ=0.263, *) | 0.005 °C/yr (τ=0.087, ns) | 0.0026189 | 0.0060837 | 0.0022539 |
| Rayong | 0.002 °C/yr (τ=-0.055, ns) | 0.005 °C/yr (τ=-0.002, ns) | 0.006 °C/yr (τ=0.159, *) | 0.000 °C/yr (τ=-0.054, ns) | 0.0033470 | 0.0039260 | -0.0014173 |
| Roi Et | 0.002 °C/yr (τ=-0.127, *) | 0.008 °C/yr (τ=0.216, *) | 0.009 °C/yr (τ=0.210, *) | 0.005 °C/yr (τ=0.160, *) | 0.0062376 | 0.0065552 | 0.0025273 |
| Sa Kaeo | 0.002 °C/yr (τ=-0.095, ns) | 0.005 °C/yr (τ=-0.000, ns) | 0.008 °C/yr (τ=0.233, *) | 0.003 °C/yr (τ=-0.021, ns) | 0.0031599 | 0.0056050 | 0.0008463 |
| Sakon Nakhon | 0.003 °C/yr (τ=-0.044, ns) | 0.008 °C/yr (τ=0.213, *) | 0.008 °C/yr (τ=0.177, *) | 0.002 °C/yr (τ=0.045, ns) | 0.0058029 | 0.0054934 | -0.0005419 |
| Samut Prakan | 0.003 °C/yr (τ=-0.000, ns) | 0.013 °C/yr (τ=0.176, *) | 0.014 °C/yr (τ=0.260, *) | 0.020 °C/yr (τ=0.457, *) | 0.0105396 | 0.0114737 | 0.0169988 |
| Samut Songkhram | 0.004 °C/yr (τ=-0.061, ns) | 0.008 °C/yr (τ=-0.156, ns) | 0.013 °C/yr (τ=-0.094, ns) | 0.004 °C/yr (τ=0.044, ns) | 0.0039526 | 0.0091715 | 0.0001221 |
| Satun | 0.001 °C/yr (τ=0.063, ns) | 0.007 °C/yr (τ=0.102, *) | 0.008 °C/yr (τ=0.173, *) | 0.006 °C/yr (τ=0.141, *) | 0.0056502 | 0.0066093 | 0.0043644 |
| Si Sa Ket | 0.004 °C/yr (τ=-0.028, ns) | 0.009 °C/yr (τ=0.232, *) | 0.011 °C/yr (τ=0.264, *) | -0.001 °C/yr (τ=-0.008, ns) | 0.0044906 | 0.0068020 | -0.0047050 |
| Songkhla | 0.001 °C/yr (τ=0.147, *) | 0.005 °C/yr (τ=0.292, *) | 0.005 °C/yr (τ=0.244, *) | 0.001 °C/yr (τ=0.081, ns) | 0.0032383 | 0.0034103 | -0.0004997 |
| Sukhothai | 0.004 °C/yr (τ=-0.018, ns) | 0.009 °C/yr (τ=0.055, ns) | 0.012 °C/yr (τ=0.095, *) | 0.009 °C/yr (τ=0.112, *) | 0.0050786 | 0.0084209 | 0.0058792 |
| Suphan Buri | 0.004 °C/yr (τ=0.043, ns) | 0.006 °C/yr (τ=0.009, ns) | 0.009 °C/yr (τ=0.225, *) | 0.002 °C/yr (τ=-0.060, ns) | 0.0015919 | 0.0052616 | -0.0024767 |
| Surat Thani | 0.005 °C/yr (τ=0.412, *) | 0.012 °C/yr (τ=0.541, *) | 0.015 °C/yr (τ=0.492, *) | 0.009 °C/yr (τ=0.490, *) | 0.0067369 | 0.0097413 | 0.0045076 |
| Surin | 0.001 °C/yr (τ=-0.166, *) | 0.002 °C/yr (τ=0.088, ns) | 0.002 °C/yr (τ=0.088, ns) | 0.002 °C/yr (τ=-0.026, ns) | 0.0012456 | 0.0016550 | 0.0016073 |
| Tak | 0.000 °C/yr (τ=-0.057, ns) | 0.002 °C/yr (τ=0.079, ns) | 0.001 °C/yr (τ=0.063, ns) | -0.001 °C/yr (τ=-0.070, ns) | 0.0013924 | 0.0010763 | -0.0015398 |
| Trang | 0.004 °C/yr (τ=0.167, *) | 0.007 °C/yr (τ=0.034, ns) | 0.010 °C/yr (τ=0.159, *) | 0.006 °C/yr (τ=0.068, ns) | 0.0022936 | 0.0055012 | 0.0014324 |
| Trat | 0.002 °C/yr (τ=0.073, ns) | 0.005 °C/yr (τ=0.108, ns) | 0.005 °C/yr (τ=0.047, ns) | 0.003 °C/yr (τ=0.059, ns) | 0.0032982 | 0.0035979 | 0.0015422 |
| Ubon Ratchathani | 0.004 °C/yr (τ=-0.061, ns) | 0.010 °C/yr (τ=0.307, *) | 0.012 °C/yr (τ=0.282, *) | 0.010 °C/yr (τ=0.206, *) | 0.0065363 | 0.0083224 | 0.0060048 |
| Udon Thani | 0.003 °C/yr (τ=-0.053, ns) | 0.012 °C/yr (τ=0.324, *) | 0.012 °C/yr (τ=0.316, *) | 0.008 °C/yr (τ=0.271, *) | 0.0090363 | 0.0094237 | 0.0048352 |
| Uthai Thani | 0.002 °C/yr (τ=0.128, ns) | 0.000 °C/yr (τ=-0.044, ns) | 0.006 °C/yr (τ=0.183, ns) | -0.007 °C/yr (τ=-0.139, ns) | -0.0018015 | 0.0040712 | -0.0085972 |
| Uttaradit | 0.005 °C/yr (τ=-0.028, ns) | 0.009 °C/yr (τ=0.271, *) | 0.012 °C/yr (τ=0.303, *) | 0.006 °C/yr (τ=0.208, *) | 0.0038231 | 0.0073585 | 0.0012578 |
| Yala | 0.002 °C/yr (τ=0.079, ns) | 0.004 °C/yr (τ=-0.021, ns) | 0.006 °C/yr (τ=0.096, ns) | 0.004 °C/yr (τ=0.011, ns) | 0.0015288 | 0.0033046 | 0.0018098 |
| Yasothon | 0.003 °C/yr (τ=0.067, ns) | 0.031 °C/yr (τ=0.300, *) | 0.030 °C/yr (τ=0.311, *) | 0.028 °C/yr (τ=0.317, *) | 0.0274395 | 0.0265341 | 0.0242895 |

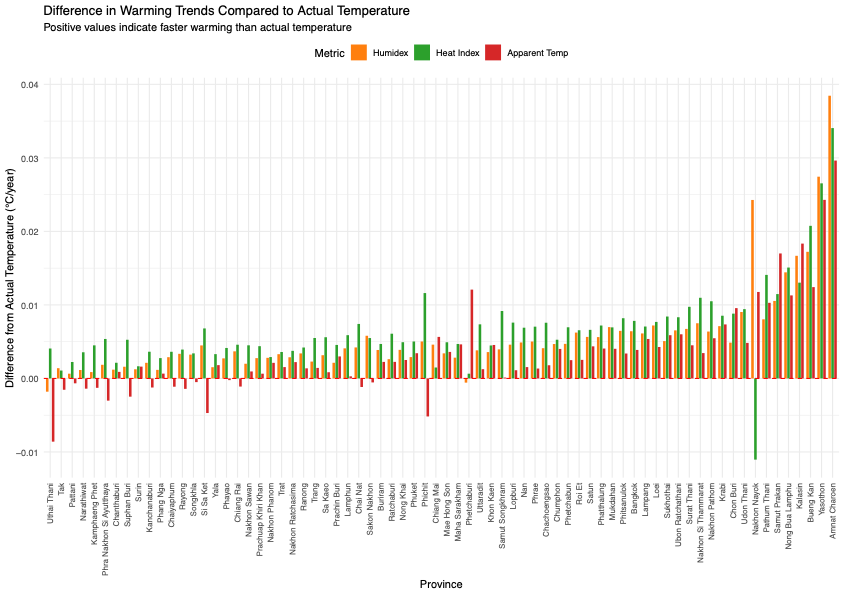

### Supplementary file S.4. Principal component analyses (clusters)

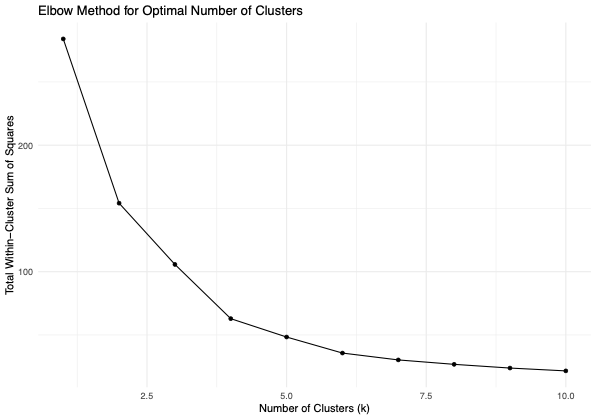

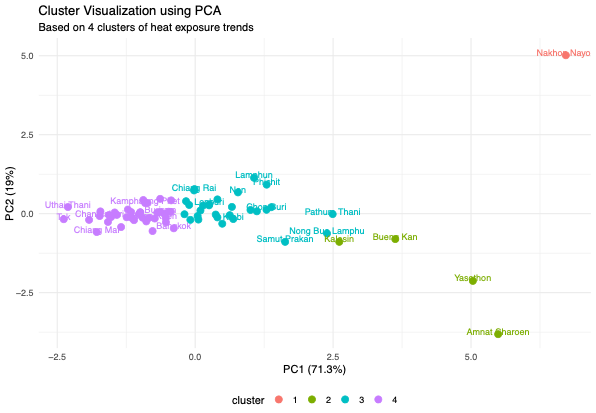

### Supplementary file S.5 Metrics summary across all provinces and years

#### Supplementary file S.5.1 Minimum, maximum, mean, median, Standard Deviation (SD)

| Metrics | Min. | Max. | Mean | Median | SD |
| --- | --- | --- | --- | --- | --- |
| Temperature | 15.66333 | 39.12581 | 28.10994 | 28.43333 | 2.150626 |
| Heat index | 16.29484 | 75.03924 | 32.45222 | 33.09621 | 4.185304 |
| Humidex | 18.35716 | 63.84101 | 38.74010 | 40.00717 | 4.265392 |
| Apparent temperature | 14.83150 | 49.78550 | 32.30949 | 33.20336 | 3.469773 |

#### Supplementary file S.5.2 Density plot

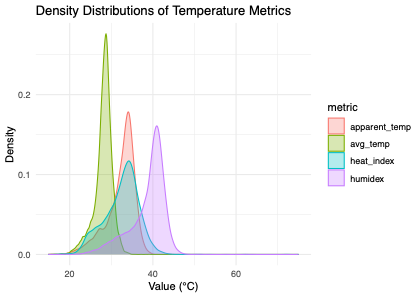

#### Supplementary file S.5.3 Distribution plot

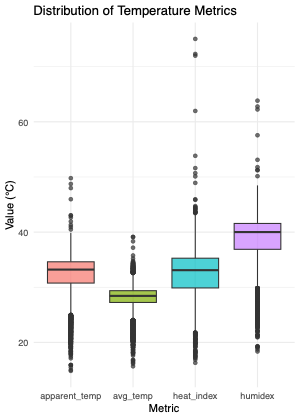

### Supplementary file S.6. Mixed effect using original heat metrics units

#### Supplementary file S.6.1. Marginal effect of total excess death explained heat metrics

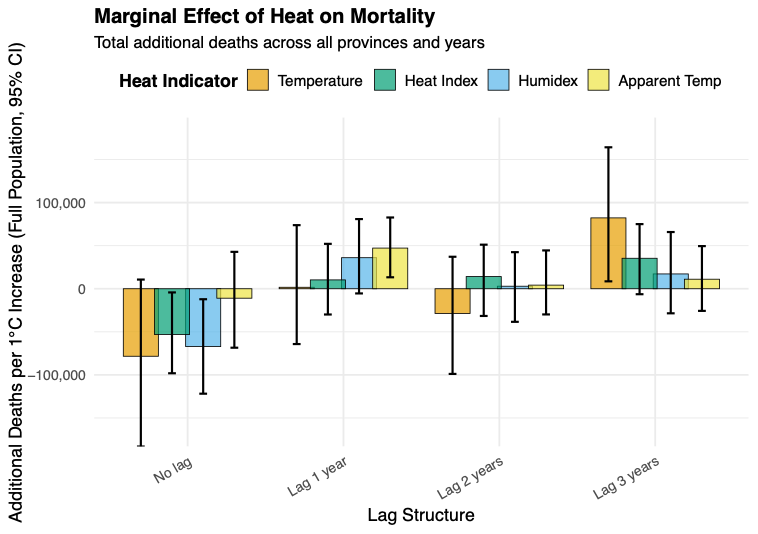

| Metrics | Lag | Total marginal effect | 95%CI - | 95%CI + |
| --- | --- | --- | --- | --- |
| Temperature | No lag | -78379.734 | -183161.56 | 10639.214 |
| Temperature | Lag 1 | 1698.14911 | -64216.899 | 73796.7051 |
| Temperature | Lag 2 | -28663.091 | -98884.804 | 37119.9368 |
| Temperature | Lag 3 | 82241.8969 | 8571.20592 | 164191.452 |
| Humidex | No lag | -67121.405 | -121837.2 | -12191.909 |
| Humidex | Lag 1 | 36036.651 | -5458.247 | 80858.5279 |
| Humidex | Lag 2 | 2930.50001 | -38416.998 | 42450.9584 |
| Humidex | Lag 3 | 17187.3802 | -28529.753 | 65820.9579 |
| Heat Index | No lag | -53131.844 | -98126.713 | -4285.9406 |
| Heat Index | Lag 1 | 10258.9964 | -29902.334 | 52109.5016 |
| Heat Index | Lag 2 | 14127.817 | -31608.037 | 51147.8179 |
| Heat Index | Lag 3 | 35358.4031 | -6360.5074 | 75003.7194 |
| Apparent Temp | No lag | -10993.588 | -68453.843 | 42824.5561 |
| Apparent Temp | Lag 1 | 47196.2888 | 13286.9543 | 82704.4393 |
| Apparent Temp | Lag 2 | 4197.81186 | -29837.964 | 44478.8961 |
| Apparent Temp | Lag 3 | 11059.6671 | -25696.321 | 49475.3604 |

#### Supplementary file S.6.2. Marginal effect of excess death per 1,000 people explained heat metrics

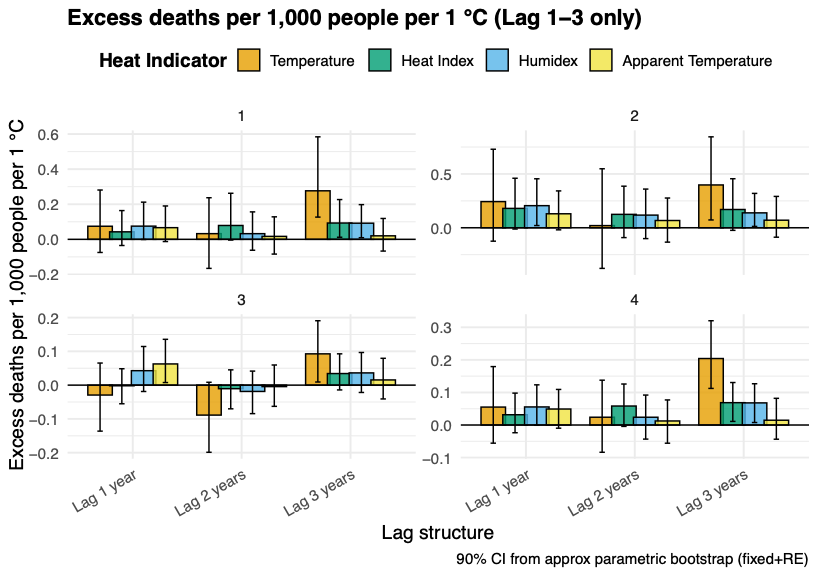

| Cluster | Metrics | Prevalence | 95%CI - | 95%CI + |
| --- | --- | --- | --- | --- |
| No lag | | | | |
| 1 | Temperature | -0.4940452 | -1.8583622 | 0.44973588 |
| 1 | Heat Index | -0.2025371 | -1.2007407 | 0.47355621 |
| 1 | Humidex | -0.3405565 | -1.4978906 | 0.2654214 |
| 1 | Apparent Temperature | -0.4508195 | -1.3286595 | 0.19903862 |
| 2 | Temperature | 0.08702903 | -0.2424995 | 0.45122812 |
| 2 | Heat Index | 0.06089127 | -0.1188413 | 0.27787747 |
| 2 | Humidex | 0.09732009 | -0.0881236 | 0.37527977 |
| 2 | Apparent Temperature | 0.12108108 | -0.0764294 | 0.34688304 |
| 3 | Temperature | -0.0850571 | -0.2013901 | 0.01489528 |
| 3 | Heat Index | -0.053431 | -0.1102132 | 0.00123109 |
| 3 | Humidex | -0.0691446 | -0.1457114 | 0.00304793 |
| 3 | Apparent Temperature | -0.0078675 | -0.0744264 | 0.06235108 |
| 4 | Temperature | -0.0930939 | -0.2121532 | 0.03899214 |
| 4 | Heat Index | -0.0729198 | -0.1373093 | -0.0156302 |
| 4 | Humidex | -0.0929693 | -0.1676897 | -0.023544 |
| 4 | Apparent Temperature | -0.0185121 | -0.0796801 | 0.04645508 |
| 1-year lag | | | | |
| 1 | Temperature | 0.07462294 | -0.0747992 | 0.28097176 |
| 1 | Heat Index | 0.0427445 | -0.0350442 | 0.1639014 |
| 1 | Humidex | 0.07520233 | -0.0008971 | 0.21192568 |
| 1 | Apparent Temperature | 0.06648306 | -0.0125646 | 0.19015381 |
| 2 | Temperature | 0.2431643 | -0.1246774 | 0.72972397 |
| 2 | Heat Index | 0.18001601 | -0.0117215 | 0.46023516 |
| 2 | Humidex | 0.20538362 | 0.02056163 | 0.45523135 |
| 2 | Apparent Temperature | 0.12990562 | -0.0191968 | 0.34255327 |
| 3 | Temperature | -0.0293568 | -0.1361623 | 0.06540648 |
| 3 | Heat Index | -0.0018647 | -0.0551518 | 0.04852022 |
| 3 | Humidex | 0.04282519 | -0.0187888 | 0.11441333 |
| 3 | Apparent Temperature | 0.06274025 | 0.00767185 | 0.13565914 |
| 4 | Temperature | 0.0550758 | -0.0557136 | 0.17932106 |
| 4 | Heat Index | 0.03154773 | -0.0237218 | 0.09787349 |
| 4 | Humidex | 0.05550035 | -0.0008315 | 0.12333957 |
| 4 | Apparent Temperature | 0.04906706 | -0.0098018 | 0.10919469 |
| 2-year lag | | | | |
| 1 | Temperature | 0.03243435 | -0.1655016 | 0.23703568 |
| 1 | Heat Index | 0.07894456 | -0.0040181 | 0.26250215 |
| 1 | Humidex | 0.032353 | -0.0621141 | 0.15644341 |
| 1 | Apparent Temperature | 0.0168406 | -0.0840454 | 0.12823799 |
| 2 | Temperature | 0.02010023 | -0.3774772 | 0.54813431 |
| 2 | Heat Index | 0.12459695 | -0.091424 | 0.38635763 |
| 2 | Humidex | 0.11753424 | -0.1007999 | 0.35956878 |
| 2 | Apparent Temperature | 0.06742613 | -0.1335991 | 0.27634739 |
| 3 | Temperature | -0.0887122 | -0.1987676 | 0.00851076 |
| 3 | Heat Index | -0.0105188 | -0.0701287 | 0.04527689 |
| 3 | Humidex | -0.0186928 | -0.0843752 | 0.04157552 |
| 3 | Apparent Temperature | -0.0043338 | -0.0627846 | 0.05961988 |
| 4 | Temperature | 0.02394 | -0.0837165 | 0.13761419 |
| 4 | Heat Index | 0.05827148 | -0.0042625 | 0.12581155 |
| 4 | Humidex | 0.02387903 | -0.0432666 | 0.09193298 |
| 4 | Apparent Temperature | 0.01242929 | -0.0558541 | 0.07696284 |
| 3-year lag | | | | |
| 1 | Temperature | 0.27653178 | 0.12663799 | 0.58406576 |
| 1 | Heat Index | 0.09297577 | 0.01191269 | 0.2265865 |
| 1 | Humidex | 0.092024 | 0.00935916 | 0.19783111 |
| 1 | Apparent Temperature | 0.01995619 | -0.066865 | 0.11888428 |
| 2 | Temperature | 0.39783965 | 0.07259381 | 0.84500385 |
| 2 | Heat Index | 0.16972041 | -0.0240506 | 0.45564927 |
| 2 | Humidex | 0.13842074 | 0.01249428 | 0.31939163 |
| 2 | Apparent Temperature | 0.07024123 | -0.0877801 | 0.29147107 |
| 3 | Temperature | 0.09261455 | 0.00925668 | 0.19067808 |
| 3 | Heat Index | 0.03431318 | -0.0141627 | 0.09266832 |
| 3 | Humidex | 0.03623251 | -0.0216849 | 0.09666379 |
| 3 | Apparent Temperature | 0.01539304 | -0.0408661 | 0.07945775 |
| 4 | Temperature | 0.20413109 | 0.11234683 | 0.32012199 |
| 4 | Heat Index | 0.06862592 | 0.01103675 | 0.1304223 |
| 4 | Humidex | 0.06792961 | 0.00760957 | 0.12665884 |
| 4 | Apparent Temperature | 0.01472918 | -0.0436287 | 0.0819407 |

#### Supplementary file S.6.3 Marginal effect of heat on Mortality (total excess deaths across all provinces and years and prevalence per 1000) – restricted to hospital deaths

#### Excess deaths

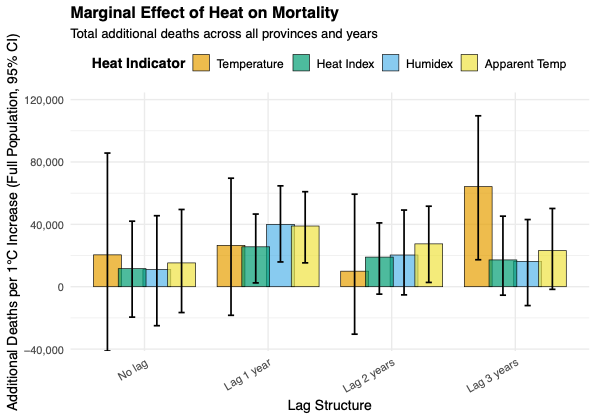

| Metrics | Lag | Predicted count | 95%CI - | 95%CI + |
| --- | --- | --- | --- | --- |
| Temperature | No lag | 20441.056 | -51402.201 | 82043.1608 |
| Temperature | Lag 1 | 26546.941 | -16817.77 | 73646.8301 |
| Temperature | Lag 2 | 9969.9875 | -28097.854 | 56413.6316 |
| Temperature | Lag 3 | 64209.3505 | 20235.3285 | 112935.537 |
| Humidex | No lag | 11035.2865 | -24318.957 | 41541.4125 |
| Humidex | Lag 1 | 39993.466 | 16302.3668 | 66404.1264 |
| Humidex | Lag 2 | 20343.1449 | -6903.6943 | 44852.3339 |
| Humidex | Lag 3 | 16285.9595 | -11573.875 | 45420.6125 |
| Heat Index | No lag | 11561.208 | -18902.343 | 40293.2141 |
| Heat Index | Lag 1 | 25598.5579 | 2570.81436 | 48810.3675 |
| Heat Index | Lag 2 | 19008.0939 | -3374.8268 | 44355.081 |
| Heat Index | Lag 3 | 17196.8805 | -8414.5699 | 41998.2918 |
| Apparent Temp | No lag | 15270.6092 | -24754.986 | 45080.8645 |
| Apparent Temp | Lag 1 | 38938.0783 | 17851.9673 | 62494.5195 |
| Apparent Temp | Lag 2 | 27481.9053 | 288.034504 | 54394.6425 |
| Apparent Temp | Lag 3 | 23206.7051 | -2053.6863 | 48195.3291 |

#### Supplementary file S.6.4 Marginal effect of excess death per 1,000 people explained heat metrics – restricted to hospital deaths

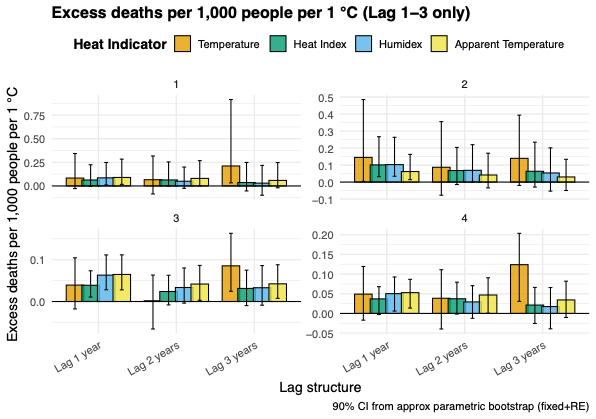

| Cluster | Metrics | Prevalence | 95%CI - | 95%CI + |
| --- | --- | --- | --- | --- |
| No lag | | | | |
| 1 | Temperature | -0.2933623 | -2.7456167 | 0.85696293 |
| 1 | Heat Index | -0.0913081 | -1.2233216 | 0.80180834 |
| 1 | Humidex | -0.0781339 | -1.0037398 | 0.50643271 |
| 1 | Apparent Temperature | -0.3970671 | -1.9893674 | 0.18581112 |
| 2 | Temperature | 0.03313835 | -0.1196018 | 0.2132111 |
| 2 | Heat Index | 0.03537337 | -0.0387846 | 0.13436182 |
| 2 | Humidex | 0.05079667 | -0.0055598 | 0.15209152 |
| 2 | Apparent Temperature | 0.04221766 | -0.0328872 | 0.15362184 |
| 3 | Temperature | 0.04297539 | -0.0260244 | 0.11708047 |
| 3 | Heat Index | 0.02646538 | -0.0103159 | 0.06617515 |
| 3 | Humidex | 0.01878404 | -0.0179443 | 0.06127356 |
| 3 | Apparent Temperature | 0.03184549 | -0.0102963 | 0.07895112 |
| 4 | Temperature | 0.00910173 | -0.0768453 | 0.07615343 |
| 4 | Heat Index | -0.0017843 | -0.0423002 | 0.04028214 |
| 4 | Humidex | -0.013859 | -0.0594912 | 0.02651914 |
| 4 | Apparent Temperature | 0.00849215 | -0.039697 | 0.04952112 |
| Lag 1 | | | | |
| 1 | Temperature | 0.08376205 | -0.0282243 | 0.34323628 |
| 1 | Heat Index | 0.06309064 | -0.0053535 | 0.22403125 |
| 1 | Humidex | 0.08576842 | 0.01056233 | 0.24854561 |
| 1 | Apparent Temperature | 0.09013832 | 0.01668754 | 0.28407909 |
| 2 | Temperature | 0.14510677 | 0.0037472 | 0.48573521 |
| 2 | Heat Index | 0.10101171 | 0.03175235 | 0.26681288 |
| 2 | Humidex | 0.10283382 | 0.03431756 | 0.26366783 |
| 2 | Apparent Temperature | 0.06191694 | 0.01417823 | 0.16306001 |
| 3 | Temperature | 0.03922032 | -0.0176924 | 0.10446462 |
| 3 | Heat Index | 0.03878027 | 0.01061381 | 0.07333901 |
| 3 | Humidex | 0.06299609 | 0.02805353 | 0.1113179 |
| 3 | Apparent Temperature | 0.06461833 | 0.0278642 | 0.11146477 |
| 4 | Temperature | 0.04904031 | -0.0170968 | 0.11922695 |
| 4 | Heat Index | 0.03693456 | -0.0028784 | 0.06787209 |
| 4 | Humidex | 0.05020941 | 0.00600997 | 0.0926456 |
| 4 | Apparent Temperature | 0.05276887 | 0.01360895 | 0.08663622 |
| Lag 2 | | | | |
| 1 | Temperature | 0.06581711 | -0.0850877 | 0.31756578 |
| 1 | Heat Index | 0.06361149 | -0.0028071 | 0.25426835 |
| 1 | Humidex | 0.0500162 | -0.025412 | 0.20010793 |
| 1 | Apparent Temperature | 0.079965 | 0.00140917 | 0.26806095 |
| 2 | Temperature | 0.08717889 | -0.0774413 | 0.35580068 |
| 2 | Heat Index | 0.06794019 | -0.0145763 | 0.20411102 |
| 2 | Humidex | 0.06906785 | -0.0002619 | 0.22001887 |
| 2 | Apparent Temperature | 0.04165623 | -0.034154 | 0.16995817 |
| 3 | Temperature | 0.00176378 | -0.0656484 | 0.06318158 |
| 3 | Heat Index | 0.0239146 | -0.0081403 | 0.06266562 |
| 3 | Humidex | 0.03344756 | -0.0038943 | 0.08014755 |
| 3 | Apparent Temperature | 0.04172898 | 0.00297885 | 0.08621932 |
| 4 | Temperature | 0.03853708 | -0.039578 | 0.11103292 |
| 4 | Heat Index | 0.03724301 | -0.0015863 | 0.07940517 |
| 4 | Humidex | 0.02928216 | -0.0123084 | 0.07051706 |
| 4 | Apparent Temperature | 0.04681456 | 0.00057482 | 0.09052392 |
| Lag 3 | | | | |
| 1 | Temperature | 0.21164499 | 0.03301715 | 0.91504869 |
| 1 | Heat Index | 0.03639348 | -0.0529558 | 0.2486986 |
| 1 | Humidex | 0.02981775 | -0.0982594 | 0.21758537 |
| 1 | Apparent Temperature | 0.05861639 | -0.0191733 | 0.24812494 |
| 2 | Temperature | 0.13964941 | -0.0195486 | 0.3939531 |
| 2 | Heat Index | 0.06349278 | -0.0294124 | 0.23490384 |
| 2 | Humidex | 0.0536622 | -0.0532738 | 0.20121646 |
| 2 | Apparent Temperature | 0.03011197 | -0.0496098 | 0.13402796 |
| 3 | Temperature | 0.08518496 | 0.02450243 | 0.162885 |
| 3 | Heat Index | 0.03125914 | -0.0097863 | 0.07488633 |
| 3 | Humidex | 0.03289044 | -0.0088391 | 0.08577781 |
| 3 | Apparent Temperature | 0.04221628 | 0.00759748 | 0.08773926 |
| 4 | Temperature | 0.12387659 | 0.03080959 | 0.20357972 |
| 4 | Heat Index | 0.02129763 | -0.0255784 | 0.06614618 |
| 4 | Humidex | 0.01745119 | -0.0391056 | 0.06564873 |
| 4 | Apparent Temperature | 0.03431023 | -0.0101992 | 0.08194119 |

#### Supplementary file S.6.5. Marginal effect of heat on Mortality (total excess deaths across all provinces and years and prevalence per 1000) – restricted to out of hospital deaths

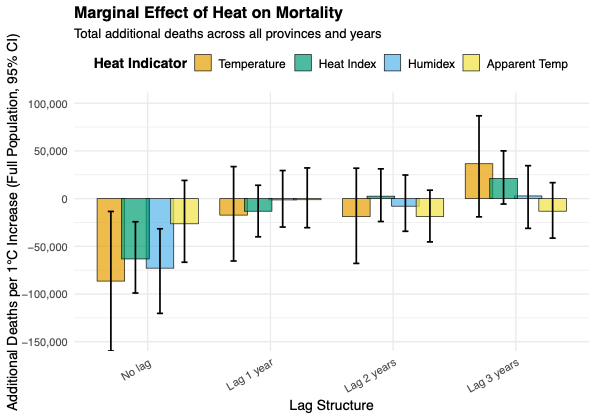

| Metrics | Lag | Predicted count | 95%CI - | 95%CI + |
| --- | --- | --- | --- | --- |
| Temperature | No lag | -86444.885 | -159732.87 | -13449.131 |
| Temperature | Lag 1 | -17280.202 | -65374.924 | 33558.8827 |
| Temperature | Lag 2 | -18838.611 | -67906.981 | 31878.1466 |
| Temperature | Lag 3 | 36539.5758 | -19095.174 | 86819.6166 |
| Humidex | No lag | -72885.945 | -120294.76 | -31588.345 |
| Humidex | Lag 1 | -1370.33 | -29788.189 | 29444.0295 |
| Humidex | Lag 2 | -7935.8765 | -34241.366 | 24731.8952 |
| Humidex | Lag 3 | 2821.39945 | -31150.137 | 34518.5349 |
| Heat Index | No lag | -63238.208 | -98931.49 | -24252.985 |
| Heat Index | Lag 1 | -13196.176 | -40025.16 | 13995.1259 |
| Heat Index | Lag 2 | 2587.92724 | -24003.825 | 31216.4284 |
| Heat Index | Lag 3 | 21201.8945 | -5687.0473 | 50037.0876 |
| Apparent Temp | No lag | -26414.971 | -66688.405 | 19083.0745 |
| Apparent Temp | Lag 1 | -1201.4556 | -30473.424 | 32178.4448 |
| Apparent Temp | Lag 2 | -18731.272 | -45370.542 | 8836.30258 |
| Apparent Temp | Lag 3 | -13172.1 | -41393.883 | 16739.5034 |

#### Supplementary file S.6.6. Marginal effect of excess death per 1,000 people explained heat metrics – restricted to out of hospital deaths

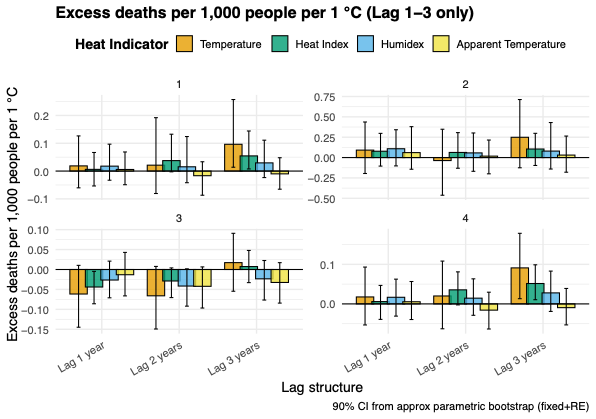

| Cluster | Metrics | Prevalence | 95%CI - | 95%CI + |
| --- | --- | --- | --- | --- |
| No lag |  |  |  |  |
| 1 | Temperature | -0.1518169 | -0.5967379 | 0.46422737 |
| 1 | Heat Index | -0.085828 | -0.7035686 | 0.31317406 |
| 1 | Humidex | -0.0781549 | -0.4465554 | 0.32489568 |
| 1 | Apparent Temperature | -0.0516029 | -0.6274084 | 0.4217 |
| 2 | Temperature | 0.13292283 | -0.1038018 | 0.47012444 |
| 2 | Heat Index | 0.06863321 | -0.0921472 | 0.26259812 |
| 2 | Humidex | 0.1005731 | -0.0018312 | 0.24699323 |
| 2 | Apparent Temperature | 0.10624282 | -0.0940411 | 0.34703006 |
| 3 | Temperature | -0.1138547 | -0.1956597 | -0.043335 |
| 3 | Heat Index | -0.0860975 | -0.1437073 | -0.0436837 |
| 3 | Humidex | -0.0782541 | -0.1260742 | -0.0428358 |
| 3 | Apparent Temperature | -0.0457906 | -0.1049624 | 0.00870413 |
| 4 | Temperature | -0.0515158 | -0.1329214 | 0.02846103 |
| 4 | Heat Index | -0.0653033 | -0.118639 | -0.0202614 |
| 4 | Humidex | -0.0550052 | -0.0982464 | -0.0185301 |
| 4 | Apparent Temperature | -0.0256586 | -0.0788891 | 0.0200836 |
| Lag 1 |  |  |  |  |
| 1 | Temperature | 0.01861811 | -0.0603063 | 0.12645203 |
| 1 | Heat Index | 0.00614387 | -0.0536201 | 0.06686656 |
| 1 | Humidex | 0.01772593 | -0.0329021 | 0.09672011 |
| 1 | Apparent Temperature | 0.00576173 | -0.0491105 | 0.06868927 |
| 2 | Temperature | 0.09154746 | -0.1955657 | 0.43677395 |
| 2 | Heat Index | 0.07782758 | -0.1046841 | 0.29725578 |
| 2 | Humidex | 0.10847572 | -0.102464 | 0.3415917 |
| 2 | Apparent Temperature | 0.06082912 | -0.143126 | 0.3795718 |
| 3 | Temperature | -0.0615437 | -0.1448586 | 0.01041346 |
| 3 | Heat Index | -0.0439692 | -0.0859978 | -0.0047217 |
| 3 | Humidex | -0.0264296 | -0.0710421 | 0.02099114 |
| 3 | Apparent Temperature | -0.0131323 | -0.0662122 | 0.0429 |
| 4 | Temperature | 0.0175913 | -0.0532635 | 0.09312127 |
| 4 | Heat Index | 0.00580534 | -0.0391135 | 0.04674419 |
| 4 | Humidex | 0.01674733 | -0.0308919 | 0.06258431 |
| 4 | Apparent Temperature | 0.00544359 | -0.0395306 | 0.05661423 |
| Lag 2 |  |  |  |  |
| 1 | Temperature | 0.02122783 | -0.0807412 | 0.19205471 |
| 1 | Heat Index | 0.03761042 | -0.0026614 | 0.13258681 |
| 1 | Humidex | 0.01536573 | -0.0418842 | 0.12420766 |
| 1 | Apparent Temperature | -0.0167023 | -0.0869236 | 0.03349899 |
| 2 | Temperature | -0.0368102 | -0.4624313 | 0.34785303 |
| 2 | Heat Index | 0.062971 | -0.1310355 | 0.3062031 |
| 2 | Humidex | 0.05736347 | -0.1685458 | 0.30199948 |
| 2 | Apparent Temperature | 0.01772314 | -0.199484 | 0.21526266 |
| 3 | Temperature | -0.0660235 | -0.1493184 | 0.00772132 |
| 3 | Heat Index | -0.0288605 | -0.0705233 | 0.00397709 |
| 3 | Humidex | -0.0415385 | -0.0919778 | 0.00165342 |
| 3 | Apparent Temperature | -0.0422076 | -0.0966857 | 0.00651336 |
| 4 | Temperature | 0.02005658 | -0.0629212 | 0.10823993 |
| 4 | Heat Index | 0.03553991 | -0.0025412 | 0.08101423 |
| 4 | Humidex | 0.01451813 | -0.0288036 | 0.06336958 |
| 4 | Apparent Temperature | -0.0157806 | -0.0634796 | 0.02958098 |
| Lag 3 |  |  |  |  |
| 1 | Temperature | 0.09637774 | 0.01382776 | 0.2573372 |
| 1 | Heat Index | 0.05456174 | 0.00760746 | 0.14418332 |
| 1 | Humidex | 0.02946552 | -0.0234141 | 0.11108552 |
| 1 | Apparent Temperature | -0.0100555 | -0.065229 | 0.04798927 |
| 2 | Temperature | 0.24897006 | -0.1253986 | 0.71255443 |
| 2 | Heat Index | 0.10507316 | -0.0970647 | 0.29775559 |
| 2 | Humidex | 0.07965369 | -0.1407217 | 0.42998993 |
| 2 | Apparent Temperature | 0.03059002 | -0.1797876 | 0.26409738 |
| 3 | Temperature | 0.01721243 | -0.0545439 | 0.0908206 |
| 3 | Heat Index | 0.00735168 | -0.0329395 | 0.04782588 |
| 3 | Humidex | -0.0233966 | -0.0766601 | 0.02255446 |
| 3 | Apparent Temperature | -0.0325015 | -0.084311 | 0.01715799 |
| 4 | Temperature | 0.09106782 | 0.01314364 | 0.17850602 |
| 4 | Heat Index | 0.05156082 | 0.01060618 | 0.0988438 |
| 4 | Humidex | 0.02784346 | -0.0190077 | 0.08294515 |
| 4 | Apparent Temperature | -0.0095017 | -0.053115 | 0.03906571 |

### Supplementary file S.7. Mixed effect using Z-scores transformed heat metrics

#### Supplementary file S.7.1. Marginal effect of total excess death explained heat metrics

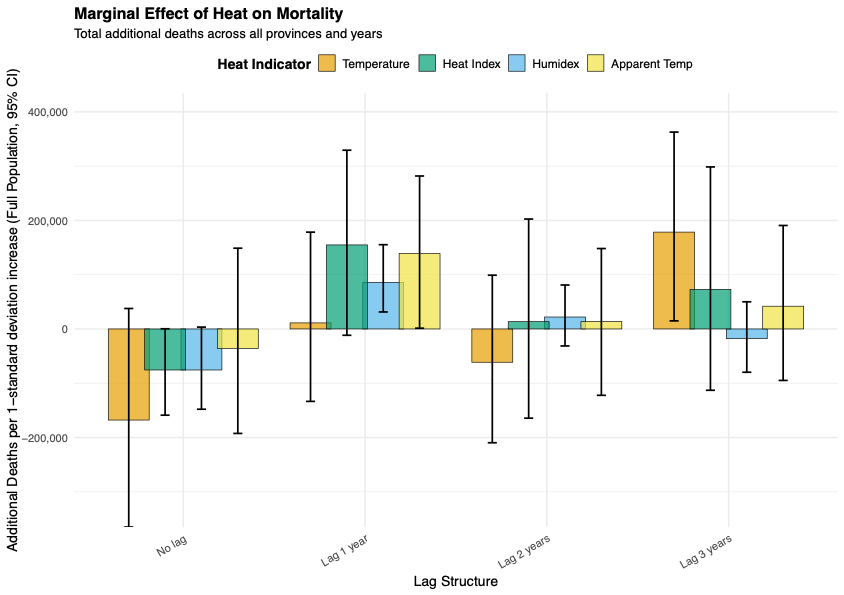

| **Metrics** | **Lag** | **Total marginal effect** | **95%CI -** | **95%CI +** |
| --- | --- | --- | --- | --- |
| Temperature | No lag | -167889.21 | -365002.44 | 37738.9789 |
| Temperature | Lag 1 year | 11171.2487 | -133462.01 | 178430.859 |
| Temperature | Lag 2 years | -61352.051 | -209703.9 | 99091.1934 |
| Temperature | Lag 3 years | 178523.794 | 14898.7184 | 362854.77 |
| Humidex | No lag | -75617.704 | -147920.62 | 3434.35234 |
| Humidex | Lag 1 year | 85865.8805 | 31333.8928 | 155285.133 |
| Humidex | Lag 2 years | 22030.8162 | -31303.564 | 80995.3906 |
| Humidex | Lag 3 years | -17720.943 | -79745.603 | 50059.008 |
| Heat Index | No lag | -75617.704 | -158898.96 | 365.562023 |
| Heat Index | Lag 1 year | 154879.93 | -11615.982 | 329428.257 |
| Heat Index | Lag 2 years | 13407.9858 | -164443.59 | 202605.09 |
| Heat Index | Lag 3 years | 72899.8598 | -112991.79 | 298625.353 |
| Apparent Temp | No lag | -36113.216 | -192543.91 | 148854.264 |
| Apparent Temp | Lag 1 year | 139141.62 | 1408.76602 | 281896.379 |
| Apparent Temp | Lag 2 years | 13730.1135 | -122263.68 | 148233.15 |
| Apparent Temp | Lag 3 years | 41946.2113 | -94843.566 | 190710 |

#### Supplementary file S.7.2. Marginal effect of excess death per 1,000 people explained heat metrics

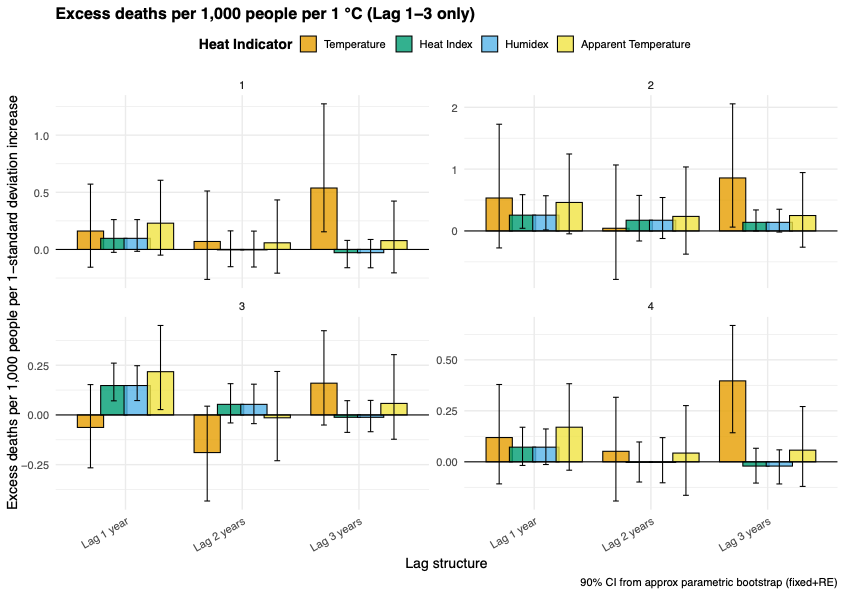

| Cluster | Metrics | Prevalence | 95%CI - | 95%CI + |
| --- | --- | --- | --- | --- |
| No lag | | | | |
| 1 | Temperature | -1.0337795 | -4.0909787 | 1.05908865 |
| 1 | Heat Index | -0.6845439 | -2.3258624 | 0.40712324 |
| 1 | Humidex | -0.6845439 | -2.2683587 | 0.38434678 |
| 1 | Apparent Temperature | -1.4982465 | -4.5666258 | 0.6791612 |
| 2 | Temperature | 0.18848666 | -0.5709834 | 0.98389206 |
| 2 | Heat Index | 0.25306407 | -0.0942481 | 0.65177992 |
| 2 | Humidex | 0.25306407 | -0.0523356 | 0.62612634 |
| 2 | Apparent Temperature | 0.42584032 | -0.2748309 | 1.39252171 |
| 3 | Temperature | -0.1815888 | -0.4147399 | 0.03594408 |
| 3 | Heat Index | -0.0570784 | -0.156991 | 0.02793004 |
| 3 | Humidex | -0.0570784 | -0.1585327 | 0.03617682 |
| 3 | Apparent Temperature | -0.0245264 | -0.2547764 | 0.17824581 |
| 4 | Temperature | -0.1987976 | -0.4615081 | 0.06233854 |
| 4 | Heat Index | -0.1330481 | -0.2289823 | -0.0418327 |
| 4 | Humidex | -0.1330481 | -0.2299631 | -0.0421049 |
| 4 | Apparent Temperature | -0.0607776 | -0.2635484 | 0.15196576 |
| Lag 1 | | | | |
| 1 | Temperature | 0.16122754 | -0.1347177 | 0.67684701 |
| 1 | Heat Index | 0.09740255 | -0.027666 | 0.24960315 |
| 1 | Humidex | 0.09740255 | -0.0302887 | 0.29216313 |
| 1 | Apparent Temperature | 0.22996842 | -0.0327341 | 0.65728667 |
| 2 | Temperature | 0.53284375 | -0.3004625 | 1.73708002 |
| 2 | Heat Index | 0.25545758 | 0.0349311 | 0.61338693 |
| 2 | Humidex | 0.25545758 | 0.04975713 | 0.61671197 |
| 2 | Apparent Temperature | 0.46105688 | -0.0235399 | 1.16382875 |
| 3 | Temperature | -0.062893 | -0.2567854 | 0.14415863 |
| 3 | Heat Index | 0.14781342 | 0.0749082 | 0.26025677 |
| 3 | Humidex | 0.14781342 | 0.06213584 | 0.2697677 |
| 3 | Apparent Temperature | 0.21746814 | -0.005674 | 0.45861876 |
| 4 | Temperature | 0.11899471 | -0.1137485 | 0.38329203 |
| 4 | Heat Index | 0.07188434 | -0.0229099 | 0.16312784 |
| 4 | Humidex | 0.07188434 | -0.0227731 | 0.17645693 |
| 4 | Apparent Temperature | 0.1697256 | -0.0257465 | 0.40115155 |
| Lag 2 | | | | |
| 1 | Temperature | 0.06987634 | -0.2708795 | 0.52274963 |
| 1 | Heat Index | -0.0036597 | -0.1524688 | 0.13584394 |
| 1 | Humidex | -0.0036597 | -0.1275504 | 0.14429207 |
| 1 | Apparent Temperature | 0.05815648 | -0.225683 | 0.4035043 |
| 2 | Temperature | 0.04330575 | -0.7118734 | 1.08024967 |
| 2 | Heat Index | 0.17319583 | -0.1421514 | 0.54170054 |
| 2 | Humidex | 0.17319583 | -0.1216527 | 0.59447415 |
| 2 | Apparent Temperature | 0.23530367 | -0.4258442 | 1.07503027 |
| 3 | Temperature | -0.1893501 | -0.41822 | 0.03058388 |
| 3 | Heat Index | 0.05328525 | -0.0470703 | 0.16527611 |
| 3 | Humidex | 0.05328525 | -0.0338452 | 0.15782807 |
| 3 | Apparent Temperature | -0.0144295 | -0.2137294 | 0.21545923 |
| 4 | Temperature | 0.05157617 | -0.1689634 | 0.29629382 |
| 4 | Heat Index | -0.002701 | -0.0921753 | 0.09259471 |
| 4 | Humidex | -0.002701 | -0.0839015 | 0.09748069 |
| 4 | Apparent Temperature | 0.04292267 | -0.1656696 | 0.26884996 |
| Lag 3 | | | | |
| 1 | Temperature | 0.53764169 | 0.17227557 | 1.18609783 |
| 1 | Heat Index | -0.0283411 | -0.150076 | 0.08394641 |
| 1 | Humidex | -0.0283411 | -0.1512806 | 0.08468326 |
| 1 | Apparent Temperature | 0.07755468 | -0.1902782 | 0.43240061 |
| 2 | Temperature | 0.85730816 | 0.03719324 | 2.27215763 |
| 2 | Heat Index | 0.13927945 | -0.0150371 | 0.34463431 |
| 2 | Humidex | 0.13927945 | -0.0146917 | 0.38912815 |
| 2 | Apparent Temperature | 0.24923843 | -0.2817404 | 0.87738786 |
| 3 | Temperature | 0.15973499 | -0.0449285 | 0.4083855 |
| 3 | Heat Index | -0.011919 | -0.0940178 | 0.06516228 |
| 3 | Humidex | -0.011919 | -0.0884009 | 0.07861603 |
| 3 | Apparent Temperature | 0.0579584 | -0.144462 | 0.28606453 |
| 4 | Temperature | 0.39683448 | 0.16171546 | 0.66084042 |
| 4 | Heat Index | -0.0209197 | -0.1011992 | 0.06730201 |
| 4 | Humidex | -0.0209197 | -0.0988317 | 0.05833532 |
| 4 | Apparent Temperature | 0.05724117 | -0.1319861 | 0.28983064 |

#### Supplementary file S.7.3 Marginal effect of heat on Mortality (total excess deaths across all provinces and years and prevalence per 1000) – restricted to hospital deaths

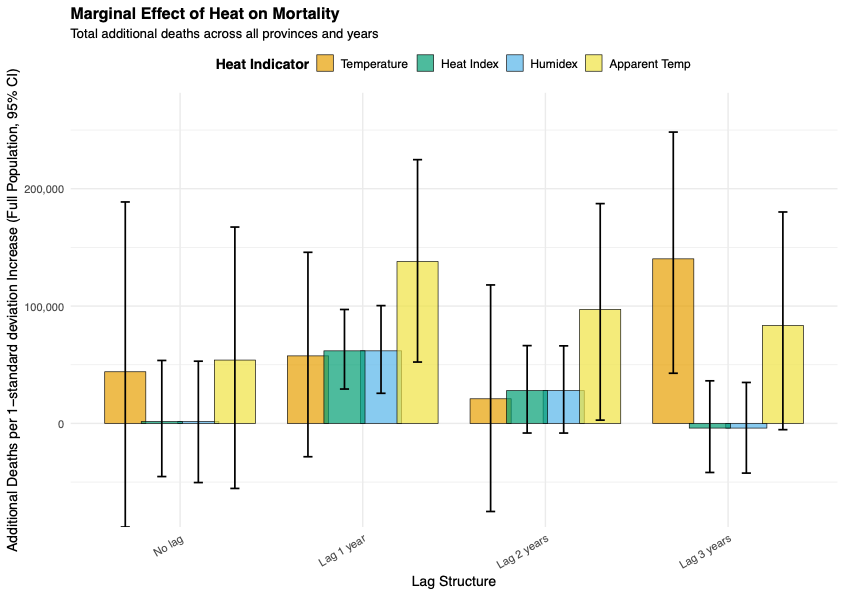

| Metrics | Lag | Total marginal effect | 95%CI - | 95%CI + |
| --- | --- | --- | --- | --- |
| Temperature | No lag | 44011.3473 | -88295.613 | 188722.061 |
| Temperature | Lag 1 year | 57550.1846 | -28404.451 | 145820.798 |
| Temperature | Lag 2 years | 20980.3208 | -75065.108 | 118000.035 |
| Temperature | Lag 3 years | 140239.248 | 42750.3649 | 248274.72 |
| Humidex | No lag | 1579.47482 | -50407.78 | 53026.765 |
| Humidex | Lag 1 year | 61843.714 | 25669.547 | 100391.626 |
| Humidex | Lag 2 years | 27991.8187 | -8224.1323 | 66097.6658 |
| Humidex | Lag 3 years | -3982.6309 | -42346.695 | 34895.1313 |
| Heat Index | No lag | 1579.47482 | -45297.881 | 53644.0177 |
| Heat Index | Lag 1 year | 61843.714 | 29292.3473 | 97078.7407 |
| Heat Index | Lag 2 years | 27991.8187 | -8192.143 | 66289.9527 |
| Heat Index | Lag 3 years | -3982.6309 | -41812.47 | 36293.8541 |
| Apparent Temp | No lag | 53990.3645 | -55421.757 | 167338.026 |
| Apparent Temp | Lag 1 year | 137946.166 | 52288.6877 | 224763.918 |
| Apparent Temp | Lag 2 years | 97127.4042 | 2796.66461 | 187316.065 |
| Apparent Temp | Lag 3 years | 83415.6178 | -5339.2693 | 180183.67 |

#### Supplementary file S.7.4 Marginal effect of excess death per 1,000 people explained heat metrics – restricted to hospital deaths

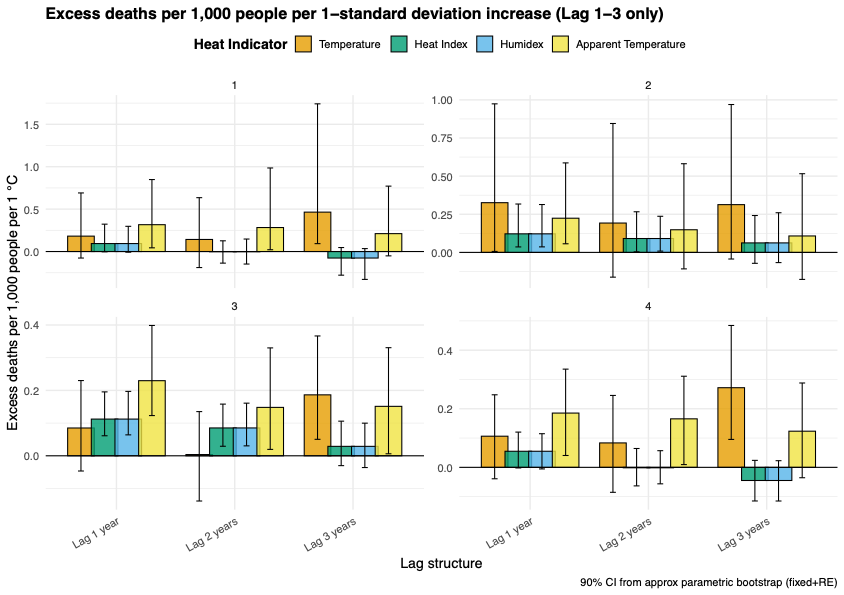

| Cluster | Metrics | Prevalence | 95%CI - | 95%CI + |
| --- | --- | --- | --- | --- |
| No lag | | | | |
| 1 | Temperature | -0.6136334 | -4.4226981 | 1.52906949 |
| 1 | Heat Index | -0.594395 | -2.4363938 | 0.52638216 |
| 1 | Humidex | -0.594395 | -3.077841 | 0.44982359 |
| 1 | Apparent Temperature | -1.2839932 | -6.2139445 | 0.85575238 |
| 2 | Temperature | 0.07203129 | -0.2610583 | 0.4443166 |
| 2 | Heat Index | 0.10481669 | 0.00364752 | 0.27428618 |
| 2 | Humidex | 0.10481669 | 0.00228359 | 0.28248447 |
| 2 | Apparent Temperature | 0.14668065 | -0.1274813 | 0.56760035 |
| 3 | Temperature | 0.09328133 | -0.0546738 | 0.2444266 |
| 3 | Heat Index | 0.03975102 | -0.020748 | 0.10937192 |
| 3 | Humidex | 0.03975102 | -0.0182656 | 0.10607675 |
| 3 | Apparent Temperature | 0.11266133 | -0.01501 | 0.28128133 |
| 4 | Temperature | 0.01964229 | -0.161491 | 0.19557109 |
| 4 | Heat Index | -0.0484813 | -0.1183803 | 0.02638563 |
| 4 | Humidex | -0.0484813 | -0.1160055 | 0.01556196 |
| 4 | Apparent Temperature | 0.0301237 | -0.1302372 | 0.22294919 |
| Lag 1 | | | | |
| 1 | Temperature | 0.18159359 | -0.0766508 | 0.69088218 |
| 1 | Heat Index | 0.09383644 | -0.0033882 | 0.32270335 |
| 1 | Humidex | 0.09383644 | -0.0077428 | 0.29744958 |
| 1 | Apparent Temperature | 0.31648357 | 0.04367948 | 0.848623 |
| 2 | Temperature | 0.32588051 | 0.00636344 | 0.97390267 |
| 2 | Heat Index | 0.12170469 | 0.03595676 | 0.31715442 |
| 2 | Humidex | 0.12170469 | 0.03620704 | 0.313872 |
| 2 | Apparent Temperature | 0.22416718 | 0.05651514 | 0.58673369 |
| 3 | Temperature | 0.08501798 | -0.0465624 | 0.23004819 |
| 3 | Heat Index | 0.11227151 | 0.06124369 | 0.19544836 |
| 3 | Humidex | 0.11227151 | 0.06400216 | 0.1969557 |
| 3 | Apparent Temperature | 0.22970809 | 0.12297594 | 0.39858271 |
| 4 | Temperature | 0.10631791 | -0.0390185 | 0.2476924 |
| 4 | Heat Index | 0.05493341 | -0.0020669 | 0.12033695 |
| 4 | Humidex | 0.05493341 | -0.0053565 | 0.11482191 |
| 4 | Apparent Temperature | 0.18527603 | 0.04048469 | 0.33528905 |
| Lag 2 | | | | |
| 1 | Temperature | 0.1424307 | -0.1889187 | 0.63505448 |
| 1 | Heat Index | -0.0026135 | -0.1366899 | 0.12648688 |
| 1 | Humidex | -0.0026135 | -0.1474014 | 0.14732905 |
| 1 | Apparent Temperature | 0.28283289 | 0.02122083 | 0.98524291 |
| 2 | Temperature | 0.19245168 | -0.1622593 | 0.8451147 |
| 2 | Heat Index | 0.09096604 | 0.0037666 | 0.26635265 |
| 2 | Humidex | 0.09096604 | 0.00840663 | 0.23683286 |
| 2 | Apparent Temperature | 0.14841593 | -0.1080446 | 0.58129127 |
| 3 | Temperature | 0.00379321 | -0.1383095 | 0.13513743 |
| 3 | Heat Index | 0.08518323 | 0.02913936 | 0.15803819 |
| 3 | Humidex | 0.08518323 | 0.03036904 | 0.16094135 |
| 3 | Apparent Temperature | 0.14802007 | 0.01958172 | 0.32994958 |
| 4 | Temperature | 0.08339569 | -0.0852125 | 0.24550867 |
| 4 | Heat Index | -0.0015301 | -0.0632799 | 0.06438634 |
| 4 | Humidex | -0.0015301 | -0.0562555 | 0.05687777 |
| 4 | Apparent Temperature | 0.16558088 | 0.00938162 | 0.31105525 |
| Lag 3 | | | | |
| 1 | Temperature | 0.46449223 | 0.09298099 | 1.74124374 |
| 1 | Heat Index | -0.0767036 | -0.2786089 | 0.04715095 |
| 1 | Humidex | -0.0767036 | -0.3285185 | 0.03462617 |
| 1 | Apparent Temperature | 0.2109073 | -0.0509799 | 0.77111843 |
| 2 | Temperature | 0.3131164 | -0.0432997 | 0.96950023 |
| 2 | Heat Index | 0.06229647 | -0.0715886 | 0.24187182 |
| 2 | Humidex | 0.06229647 | -0.0668587 | 0.26004199 |
| 2 | Apparent Temperature | 0.10776502 | -0.1769875 | 0.51592079 |
| 3 | Temperature | 0.18638424 | 0.05039085 | 0.36648116 |
| 3 | Heat Index | 0.02887181 | -0.0301321 | 0.10588485 |
| 3 | Humidex | 0.02887181 | -0.0359645 | 0.09978509 |
| 3 | Apparent Temperature | 0.15123142 | 0.0061215 | 0.33060476 |
| 4 | Temperature | 0.27186899 | 0.09534194 | 0.48435026 |
| 4 | Heat Index | -0.0448992 | -0.1148153 | 0.02383887 |
| 4 | Humidex | -0.0448992 | -0.1149114 | 0.02274239 |
| 4 | Apparent Temperature | 0.12345088 | -0.0354865 | 0.28796248 |

#### Supplementary file S.7.5. Marginal effect of heat on Mortality (total excess deaths across all provinces and years and prevalence per 1000) – restricted to out of hospital deaths

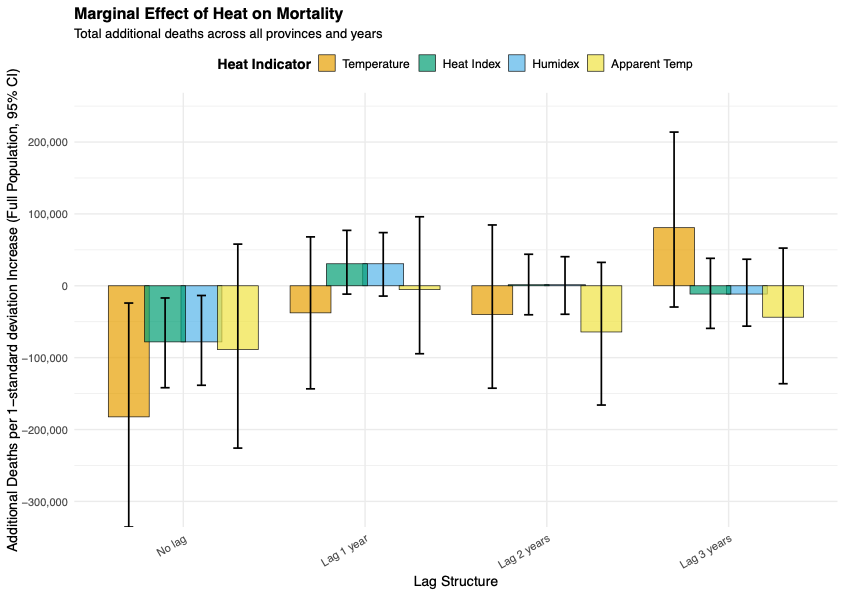

| Metrics | Lag | Total marginal effect | 95%CI - | 95%CI + |
| --- | --- | --- | --- | --- |
| Temperature | No lag | -182375.49 | -335610.67 | -24000.454 |
| Temperature | Lag 1 year | -37681.277 | -143373.16 | 68043.0757 |
| Temperature | Lag 2 years | -40030.302 | -142480.1 | 84630.0463 |
| Temperature | Lag 3 years | 80811.911 | -29542.896 | 213855.612 |
| Humidex | No lag | -78065.244 | -138406.03 | -13506.325 |
| Humidex | Lag 1 year | 30622.94 | -14312.77 | 73993.2822 |
| Humidex | Lag 2 years | 1448.18822 | -39609.115 | 40489.8819 |
| Humidex | Lag 3 years | -11510.065 | -56145.127 | 36944.1934 |
| Heat Index | No lag | -78065.244 | -141815.1 | -16941.783 |
| Heat Index | Lag 1 year | 30622.94 | -11576.177 | 77097.955 |
| Heat Index | Lag 2 years | 1448.18822 | -40330.246 | 43860.5454 |
| Heat Index | Lag 3 years | -11510.065 | -59258.057 | 38215.3221 |
| Apparent Temp | No lag | -88663.113 | -225813.93 | 57974.0407 |
| Apparent Temp | Lag 1 year | -5245.5738 | -94524.01 | 96026.9965 |
| Apparent Temp | Lag 2 years | -64251.992 | -165912.8 | 32560.5726 |
| Apparent Temp | Lag 3 years | -43682.935 | -136238.99 | 52453.2147 |

#### Supplementary file S.7.6. Marginal effect of excess death per 1,000 people explained heat metrics – restricted to out of hospital deaths

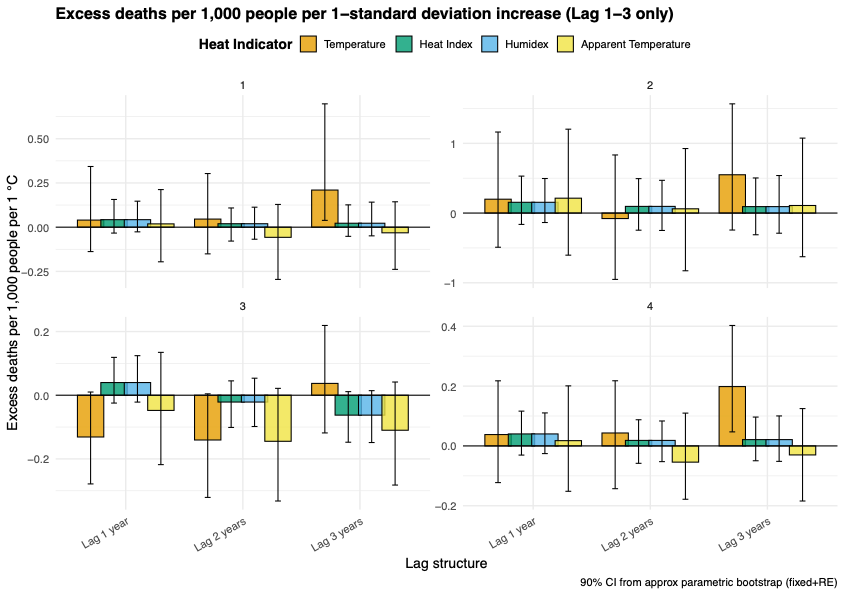

| Cluster | Metrics | Prevalence | 95%CI - | 95%CI + |
| --- | --- | --- | --- | --- |
| No lag | | | | |
| 1 | Temperature | -0.336252 | -1.751668 | 0.89720256 |
| 1 | Heat Index | -0.08834 | -0.931623 | 0.69837982 |
| 1 | Humidex | -0.08834 | -0.8410617 | 0.65427186 |
| 1 | Apparent Temperature | -0.1792546 | -1.9228665 | 1.37067298 |
| 2 | Temperature | 0.25558577 | -0.4269798 | 1.16027349 |
| 2 | Heat Index | 0.19907183 | -0.1734984 | 0.64222034 |
| 2 | Humidex | 0.19907183 | -0.1632299 | 0.62954281 |
| 2 | Apparent Temperature | 0.38264966 | -0.4109403 | 1.42570245 |
| 3 | Temperature | -0.275894 | -0.4706782 | -0.1179274 |
| 3 | Heat Index | -0.106746 | -0.1983369 | -0.0330972 |
| 3 | Humidex | -0.106746 | -0.1962696 | -0.0351909 |
| 3 | Apparent Temperature | -0.1549714 | -0.3429739 | 0.03692854 |
| 4 | Temperature | -0.1593323 | -0.3541669 | 0.00231039 |
| 4 | Heat Index | -0.0903998 | -0.1765666 | -0.0123651 |
| 4 | Humidex | -0.0903998 | -0.1830984 | -0.0190174 |
| 4 | Apparent Temperature | -0.0858894 | -0.2578016 | 0.10547461 |
| Lag 1 | | | | |
| 1 | Temperature | 0.04014707 | -0.1379293 | 0.34306428 |
| 1 | Heat Index | 0.04242683 | -0.0333493 | 0.15674706 |
| 1 | Humidex | 0.04242683 | -0.0264196 | 0.14694547 |
| 1 | Apparent Temperature | 0.01865006 | -0.1957453 | 0.21246509 |
| 2 | Temperature | 0.19874615 | -0.4901018 | 1.16207984 |
| 2 | Heat Index | 0.15449027 | -0.1627011 | 0.52997089 |
| 2 | Humidex | 0.15449027 | -0.1356766 | 0.49554321 |
| 2 | Apparent Temperature | 0.21339134 | -0.6037116 | 1.20294592 |
| 3 | Temperature | -0.1311802 | -0.2785831 | 0.01001045 |
| 3 | Heat Index | 0.03976837 | -0.0246369 | 0.11889662 |
| 3 | Humidex | 0.03976837 | -0.0215609 | 0.12421799 |
| 3 | Apparent Temperature | -0.0477336 | -0.217996 | 0.13462114 |
| 4 | Temperature | 0.03793291 | -0.1226462 | 0.21756372 |
| 4 | Heat Index | 0.04008079 | -0.0306136 | 0.1162801 |
| 4 | Humidex | 0.04008079 | -0.0256849 | 0.11028093 |
| 4 | Apparent Temperature | 0.01762028 | -0.1516507 | 0.2007198 |
| Lag 2 | | | | |
| 1 | Temperature | 0.04576924 | -0.1508795 | 0.30315672 |
| 1 | Heat Index | 0.01952002 | -0.0785818 | 0.10862205 |
| 1 | Humidex | 0.01952002 | -0.0681085 | 0.11300381 |
| 1 | Apparent Temperature | -0.0573962 | -0.295218 | 0.12860038 |
| 2 | Temperature | -0.078882 | -0.9513153 | 0.83322063 |
| 2 | Heat Index | 0.09567097 | -0.2456091 | 0.49382081 |
| 2 | Humidex | 0.09567097 | -0.2494955 | 0.46954496 |
| 2 | Apparent Temperature | 0.06101606 | -0.827533 | 0.92519805 |
| 3 | Temperature | -0.1406366 | -0.3209262 | 0.00431164 |
| 3 | Heat Index | -0.0214382 | -0.1011902 | 0.04507518 |
| 3 | Humidex | -0.0214382 | -0.0982786 | 0.0534471 |
| 3 | Apparent Temperature | -0.1446711 | -0.332247 | 0.02157884 |
| 4 | Temperature | 0.04324391 | -0.1429959 | 0.21774879 |
| 4 | Heat Index | 0.0184426 | -0.0581632 | 0.08746368 |
| 4 | Humidex | 0.0184426 | -0.0525358 | 0.08333981 |
| 4 | Apparent Temperature | -0.0542294 | -0.1782374 | 0.10963833 |
| Lag 3 | | | | |
| 1 | Temperature | 0.20987797 | 0.03847026 | 0.69718726 |
| 1 | Heat Index | 0.02231783 | -0.0517782 | 0.12615454 |
| 1 | Humidex | 0.02231783 | -0.0490298 | 0.14154382 |
| 1 | Apparent Temperature | -0.0318774 | -0.2384832 | 0.1434922 |
| 2 | Temperature | 0.54929416 | -0.2435056 | 1.56625869 |
| 2 | Heat Index | 0.09241763 | -0.3111241 | 0.50347801 |
| 2 | Humidex | 0.09241763 | -0.2889651 | 0.5382551 |
| 2 | Apparent Temperature | 0.10852149 | -0.6260864 | 1.07488542 |
| 3 | Temperature | 0.03710826 | -0.1184432 | 0.21910069 |
| 3 | Heat Index | -0.0625662 | -0.1476062 | 0.01123772 |
| 3 | Humidex | -0.0625662 | -0.1485988 | 0.01432164 |
| 3 | Apparent Temperature | -0.1099811 | -0.2821626 | 0.04147441 |
| 4 | Temperature | 0.19831477 | 0.04691867 | 0.40259599 |
| 4 | Heat Index | 0.02108931 | -0.0496194 | 0.09642927 |
| 4 | Humidex | 0.02108931 | -0.0516013 | 0.10020953 |
| 4 | Apparent Temperature | -0.0301222 | -0.1841368 | 0.12475254 |

### Supplementary file S.8. Main models specification

| **Original health metrics units** | | | | | | | |
| --- | --- | --- | --- | --- | --- | --- | --- |
| **Outcome** | **Exposure** | **Lag** | **AIC** | **BIC** | **LogLik** | **Deviance** | **DF_Resid** |
| Total Deaths (No Cluster) | Temperature | No Lag | 13903.0949 | 14000.8867 | -6931.5474 | 880.114387 | 962 |
| Total Deaths (No Cluster) | Humidex | No Lag | 13883.2878 | 13981.0796 | -6921.6439 | 872.339758 | 962 |
| Total Deaths (No Cluster) | Heat Index | No Lag | 13885.7472 | 13983.539 | -6922.8736 | 872.802548 | 962 |
| Total Deaths (No Cluster) | Apparent Temp | No Lag | 13875.9007 | 13973.6925 | -6917.9503 | 876.873897 | 962 |
| Total Deaths (No Cluster) | Temperature | Lag1 | 10938.6542 | 11017.6208 | -5452.3271 | 730.225336 | 752 |
| Total Deaths (No Cluster) | Humidex | Lag1 | 10921.6005 | 11000.5671 | -5443.8003 | 687.990533 | 752 |
| Total Deaths (No Cluster) | Heat Index | Lag1 | 10925.653 | 11004.6196 | -5445.8265 | 684.634506 | 752 |
| Total Deaths (No Cluster) | Apparent Temp | Lag1 | 10926.4228 | 11005.3894 | -5446.2114 | 720.613497 | 752 |
| Total Deaths (No Cluster) | Temperature | Lag2 | 10924.3344 | 11003.301 | -5445.1672 | 678.526391 | 752 |
| Total Deaths (No Cluster) | Humidex | Lag2 | 10917.5119 | 10996.4785 | -5441.756 | 679.579477 | 752 |
| Total Deaths (No Cluster) | Heat Index | Lag2 | 10910.7617 | 10989.7283 | -5438.3809 | 676.268976 | 752 |
| Total Deaths (No Cluster) | Apparent Temp | Lag2 | 10924.9707 | 11003.9372 | -5445.4853 | 682.468287 | 752 |
| Total Deaths (No Cluster) | Temperature | Lag3 | 10912.4954 | 10991.462 | -5439.2477 | 677.272553 | 752 |
| Total Deaths (No Cluster) | Humidex | Lag3 | 10904.4827 | 10983.4492 | -5435.2413 | 671.323053 | 752 |
| Total Deaths (No Cluster) | Heat Index | Lag3 | 10907.7056 | 10986.6721 | -5436.8528 | 674.385138 | 752 |
| Total Deaths (No Cluster) | Apparent Temp | Lag3 | 10924.4471 | 11003.4137 | -5445.2236 | 683.194881 | 752 |
| Total Deaths (With Cluster) | Temperature | No Lag | 13233.2852 | 13360.4146 | -6590.6426 | 899.247716 | 956 |
| Total Deaths (With Cluster) | Humidex | No Lag | 13230.0701 | 13357.1995 | -6589.0351 | 902.066764 | 956 |
| Total Deaths (With Cluster) | Heat Index | No Lag | 13226.8215 | 13353.9509 | -6587.4107 | 893.927828 | 956 |
| Total Deaths (With Cluster) | Apparent Temp | No Lag | 13224.7782 | 13351.9076 | -6586.3891 | 890.126445 | 956 |
| Total Deaths (With Cluster) | Temperature | Lag1 | 10247.6927 | 10349.8847 | -5101.8463 | 712.180354 | 747 |
| Total Deaths (With Cluster) | Humidex | Lag1 | 10233.8726 | 10336.0646 | -5094.9363 | 713.014125 | 747 |
| Total Deaths (With Cluster) | Heat Index | Lag1 | 10239.9369 | 10342.1289 | -5097.9685 | 712.77078 | 747 |
| Total Deaths (With Cluster) | Apparent Temp | Lag1 | 10235.3042 | 10337.4962 | -5095.6521 | 712.121972 | 747 |
| Total Deaths (With Cluster) | Temperature | Lag2 | 10246.161 | 10348.353 | -5101.0805 | 697.576005 | 747 |
| Total Deaths (With Cluster) | Humidex | Lag2 | 10240.4971 | 10342.6891 | -5098.2486 | 700.462099 | 747 |
| Total Deaths (With Cluster) | Heat Index | Lag2 | 10237.3275 | 10339.5195 | -5096.6638 | 694.530514 | 747 |
| Total Deaths (With Cluster) | Apparent Temp | Lag2 | 10234.7884 | 10336.9804 | -5095.3942 | 693.131464 | 747 |
| Total Deaths (With Cluster) | Temperature | Lag3 | 10222.4572 | 10324.6492 | -5089.2286 | 687.716259 | 747 |
| Total Deaths (With Cluster) | Humidex | Lag3 | 10227.824 | 10330.016 | -5091.912 | 677.108115 | 747 |
| Total Deaths (With Cluster) | Heat Index | Lag3 | 10222.0864 | 10324.2784 | -5089.0432 | 675.40141 | 747 |
| Total Deaths (With Cluster) | Apparent Temp | Lag3 | 10237.7084 | 10339.9004 | -5096.8542 | 687.400029 | 747 |
| **Z-scores transformed heat metrics** | | | | | | | |
| Total Deaths (No Cluster) | Temperature | No Lag | 13903.0948 | 14000.8866 | -6931.5474 | 880.023464 | 962 |
| Total Deaths (No Cluster) | Humidex | No Lag | 13894.9347 | 13992.7265 | -6927.4674 | 881.69688 | 962 |
| Total Deaths (No Cluster) | Heat Index | No Lag | 13894.9347 | 13992.7265 | -6927.4674 | 881.69688 | 962 |
| Total Deaths (No Cluster) | Apparent Temp | No Lag | 13876.3502 | 13974.142 | -6918.1751 | 877.010694 | 962 |
| Total Deaths (No Cluster) | Temperature | Lag1 | 10931.7137 | 11010.6802 | -5448.8568 | 685.736571 | 752 |
| Total Deaths (No Cluster) | Humidex | Lag1 | 10913.9796 | 10992.9461 | -5439.9898 | 694.216996 | 752 |
| Total Deaths (No Cluster) | Heat Index | Lag1 | 10921.6004 | 11000.567 | -5443.8002 | 688.06794 | 752 |
| Total Deaths (No Cluster) | Apparent Temp | Lag1 | 10920.6921 | 10999.6587 | -5443.3461 | 689.625527 | 752 |
| Total Deaths (No Cluster) | Temperature | Lag2 | 10924.3344 | 11003.3009 | -5445.1672 | 678.228372 | 752 |
| Total Deaths (No Cluster) | Humidex | Lag2 | 10916.7715 | 10995.738 | -5441.3857 | 683.294732 | 752 |
| Total Deaths (No Cluster) | Heat Index | Lag2 | 10917.5118 | 10996.4783 | -5441.7559 | 679.564914 | 752 |
| Total Deaths (No Cluster) | Apparent Temp | Lag2 | 10925.0748 | 11004.0414 | -5445.5374 | 682.356972 | 752 |
| Total Deaths (No Cluster) | Temperature | Lag3 | 10912.4954 | 10991.4619 | -5439.2477 | 677.103879 | 752 |
| Total Deaths (No Cluster) | Humidex | Lag3 | 10902.6003 | 10981.5669 | -5434.3002 | 673.021225 | 752 |
| Total Deaths (No Cluster) | Heat Index | Lag3 | 10904.4825 | 10983.4491 | -5435.2413 | 671.024364 | 752 |
| Total Deaths (No Cluster) | Apparent Temp | Lag3 | 10924.3948 | 11003.3613 | -5445.1974 | 683.338091 | 752 |
| Total Deaths (With Cluster) | Temperature | No Lag | 13912.5879 | 14039.7172 | -6930.2939 | 880.736459 | 956 |
| Total Deaths (With Cluster) | Humidex | No Lag | 13900.9471 | 14028.0765 | -6924.4736 | 884.917231 | 956 |
| Total Deaths (With Cluster) | Heat Index | No Lag | 13900.9471 | 14028.0765 | -6924.4736 | 884.917231 | 956 |
| Total Deaths (With Cluster) | Apparent Temp | No Lag | 13884.3337 | 14011.463 | -6916.1668 | 878.677132 | 956 |
| Total Deaths (With Cluster) | Temperature | Lag1 | 10937.6849 | 11039.8769 | -5446.8424 | 686.649294 | 747 |
| Total Deaths (With Cluster) | Humidex | Lag1 | 10917.8262 | 11020.0182 | -5436.9131 | 698.39882 | 747 |
| Total Deaths (With Cluster) | Heat Index | Lag1 | 10917.8262 | 11020.0182 | -5436.9131 | 698.39882 | 747 |
| Total Deaths (With Cluster) | Apparent Temp | Lag1 | 10926.9458 | 11029.1378 | -5441.4729 | 692.725538 | 747 |
| Total Deaths (With Cluster) | Temperature | Lag2 | 10930.1838 | 11032.3758 | -5443.0919 | 678.620241 | 747 |
| Total Deaths (With Cluster) | Humidex | Lag2 | 10923.4813 | 11025.6733 | -5439.7406 | 685.629346 | 747 |
| Total Deaths (With Cluster) | Heat Index | Lag2 | 10923.4813 | 11025.6733 | -5439.7406 | 685.629346 | 747 |
| Total Deaths (With Cluster) | Apparent Temp | Lag2 | 10933.0865 | 11035.2785 | -5444.5432 | 682.67373 | 747 |
| Total Deaths (With Cluster) | Temperature | Lag3 | 10917.3283 | 11019.5203 | -5436.6641 | 680.496237 | 747 |
| Total Deaths (With Cluster) | Humidex | Lag3 | 10928.3276 | 11030.5196 | -5442.1638 | 703.004588 | 747 |
| Total Deaths (With Cluster) | Heat Index | Lag3 | 10928.3276 | 11030.5196 | -5442.1638 | 703.004588 | 747 |
| Total Deaths (With Cluster) | Apparent Temp | Lag3 | 10931.9431 | 11034.1351 | -5443.9716 | 684.428978 | 747 |
